# Can leptin-specific epigenetic modulation of preterm cord blood predispose obesity?

**DOI:** 10.1101/2024.12.16.24319077

**Authors:** Navya Sree Boga, Amit K Banerjee, Saikanth Varma, Archana Molangiri, Syeda Farhana, Santosh Kumar Banjara, Nitasha Bagga, Asim K. Duttaroy, Sanjay Basak

**Affiliations:** National Institute of Nutrition, Indian Council of Medical Research, Hyderabad, India; Rainbow Children’s Hospital & Birth Right, Banjara hills, Hyderabad, India; Department of Nutrition, Institute of Basic Medical Sciences, Faculty of Medicine, University of Oslo, Norway

**Keywords:** Preterm, Obesity, Leptin, Methylation, MicroRNA

## Abstract

**Objective:** This retrospective cohort investigated the role of leptin’s promoter methylation and microRNA targeting profile in developing adiposity and inflammation in neonates, using umbilical cord blood from preterm (n=67) and term (n=71) mothers.

**Methods:** Global DNA methylation and leptin promoter methylation were performed. ELISA determined leptin and IGF1 levels. Real-time PCR measured mRNA levels. MicroRNA target prediction on the human leptin gene (*LEP*) was done *in silico* using network analysis.

**Results:** Preterm cord blood significantly reduced genome-wide (p<0.001) and *LEP* promoter methylation (p=0.001), increased *LEP* & *LEPR* expression (p=0.04), and circulatory leptin (p=0.41). Neonatal birth weight positively correlated with leptin and IGF1 levels in preterm (r=0.47, p=0.04) but not in the term. *IL6* expression showed a positive correlation with circulatory leptin (r= 0.687, p=0.008), *LEP* (r= 0.763, p=0.009), and an inverse association with *LEP* promoter methylation (r= -0.636, p=0.04) in preterm. The obtained *LEP* targeting miRNAs showed their affinities for critical genes associated with body fat distribution, fat cell differentiation, and energy regulation, implicating a close association in the *LEP*-miRNA-obesity axis.

**Conclusions:** The strong correlation between *LEP* methylation and pro-inflammatory cytokine influences each other in developing chronic inflammation in preterm neonates, which might predispose them to obesity in later life.

**Study importance:** **What is already known?**

- Leptin communicates about the body’s fat deposits to the brain and aids in maintaining energy homeostasis and stable body weight.
- Preterm exhibit lower body weight and fat mass at birth than term neonates, who often show rapid compensatory catch-up growth.

**What does this study add?**

- Leptin gene (*LEP*) promoter methylation was reduced in preterm cord blood compared to term.
- Higher interleukin-6 (*IL6*) and tumour necrosis factor-alpha (*TNF*_α_) expression in preterm but not in term. *IL6* correlated positively with circulatory leptin and *LEP* expression while inversely associated with *LEP*-specific promoter methylation, indicating that a dysregulated epigenetic control can promote low-grade inflammation in preterm neonates.
- *LEP*-targeting micro-RNAs showed affinities for critical genes associated with fat cell differentiation, energy regulation, and other processes.

**How might these results change the direction of research or the focus of clinical practice?**

- Since others observed dysregulated *LEP* methylation in the adipose tissue of obese subjects, these data imply that leptin could mediate the risk for obesity during preterm birth.
- While short-term outcomes of preterm birth are well addressed, its effect on long-term metabolic health is of concern as it might elevate the risk of obesity.

**Graphical Abstract:** Maternal factors leading to preterm birth and cord blood leptin dysregulation in predicting obesity. Elevated blood pressure, infection, and lower haemoglobin in preterm disrupted epigenetic control of leptin and activated inflammation that might induce leptin resistance. The latter is known to reduce satiety and increase body mass, elevating the risk of obesity. Solid arrows depict present data, and dotted lines indicate possible pathways.

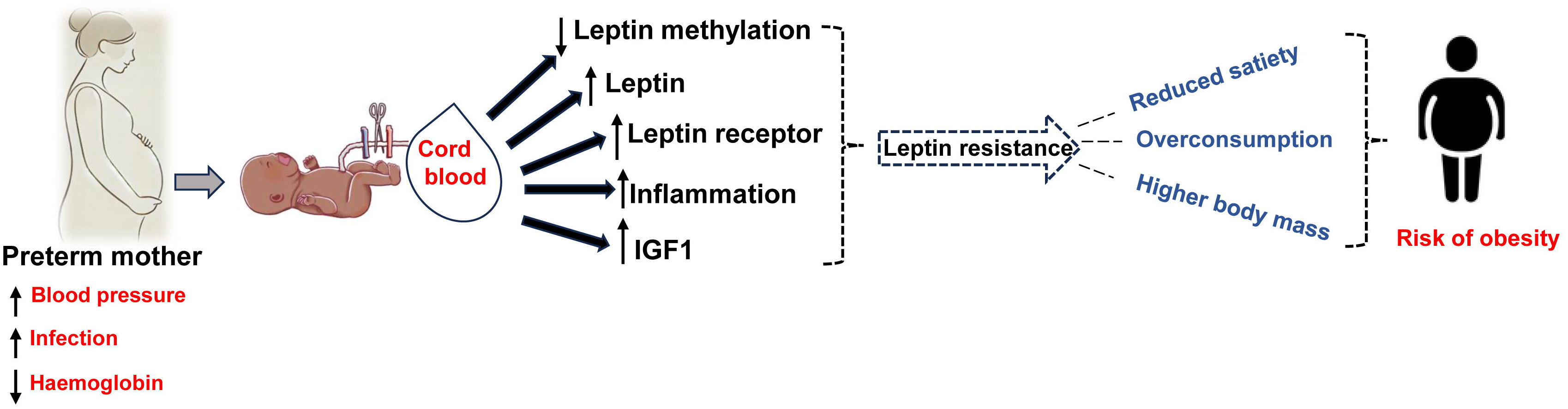

## 1. Introduction

Leptin is a well-known adipokine and is produced by the white adipose tissues where the expression of the leptin gene (*LEP*) and its release remains proportional to the fat deposition (1), induces pro-adipogenic and proinflammatory signalling in adipocytes (2), and a key regulator of the adipose organ (3). Due to the rapid rise in obesity and its long-term consequences, preterm birth has gained increased focus *in utero* programming of obesity. Lower leptin in preterm neonates is a protective response to boost the hunger for catch-up growth (4). However, low birth weight adults showed higher plasma leptin retained during catch-up growth and later life (5). The reasons for leptin resistance are unknown, however, impaired preadipocyte maturation, endocrine regulation, inflammation, nutrition, and epigenetics could be involved. Preterm neonates often show rapid compensatory catch-up growth, which might lead to obesity in later life (6). Although leptin modulates adiposity, its epigenetic role in the predictive risk of neonates’ later obesity in preterm is unknown.

*LEP* methylation refers to adding a methyl group to the leptin gene, which can affect its expression. Preterm birth, being an early-life stressor, showed altered global methylation (7), indicating the role of epigenetic factors on fetal growth. Increasing evidence suggests an association between leptin and an increased risk of inflammatory diseases (8) since serum leptin is elevated in chronic inflammatory conditions. Elevated inflammation can impair the leptin signalling pathway by interrupting cellular functions (2). Although leptin expression and level are lower in preterm, their roles in leptin promoter methylation concerning inflammation are not reported in preterm.

Many reports mentioned the active and passive role of the *LEP* gene expression in regulating adipocyte differentiation (1, 9) and lipid metabolism (10, 11). However, without knowing the plausible link between the mRNA’s expression with neonatal adiposity and miRNAs that can potentially target and regulate the expression of the *LEP* gene, the predictive risks of developing obesity due to epigenetic alteration are not possible. Among various markers for obesity, leptin is a well-established biomarker, and its expression is correlated with the degree of adiposity (12). This adiposity is further regulated by multiple miRNAs associated with leptin and related genes that target food intake, influence the hypothalamic leptin sensitivity (13), thermal regulation, and metabolic conditions of the body, and play a crucial part in the onset and progress of obesity (14). MicroRNAs are well-known epigenetic controllers of housekeeping and other gene expression and are often associated with various disease conditions, including obesity (15). For example, miRNA-221, present in human adipose tissue, was reported to be upregulated in obesity, influencing fat metabolism while acting on the downstream of the *LEP* gene (16). In two independent cohorts, specific miRNAs such as hsa-miR-532-5p were positively associated with neonates’ birth weight (17). However, no studies have attempted to predict the most susceptible targeting miRNAs for the human *LEP* to find a plausible link between miRNA expression and neonatal adiposity risk and miRNAs that can potentially target and regulate the expression of the *LEP* gene.

The study examined the role of epigenetic modification of the leptin promoter in modulating adiposity factors and its predictive risk of neonates’ later obesity using umbilical cord blood from preterm and term subjects. In addition, it examined the regulation of neonatal leptin methylation by maternal factors.

## 2. Materials and methods

### 2.1 Clinical subjects and samples

The term (n=71) and preterm (n=67) subjects were categorized based on gestational age (GA), and birth weight. The umbilical cord blood was collected from these subjects at Rainbow Children’s Hospital & Birthright, Hyderabad, India. Cord blood collection was done following the principles of the Declaration of Helsinki. This retrospective cohort was approved by the Institutional Ethical Committee of the ICMR-National Institute of Nutrition (EC/NEW/INST/2021/1206) and the Ethics Committee of Rainbow Children’s Medicare (EC/LMS1/Sept-23/15). Mothers were informed about the objectives of the study and provided consent for participation. Subjects with congenital anomalies, familial metabolic disorders, and previous preterm history were excluded.

### 2.2 Cord blood collection

The umbilical cord venous blood was drawn in EDTA-coated vacutainers and clot-activator vacutainers to separate plasma and serum respectively. Plasma and serum were stored at -80°C for further analysis.

### 2.3 Serum leptin and IGF1 estimation

Circulating levels of leptin and IGF1 were measured by the ELISA kits (#H6017, Elabscience) and (#DY291-05, R&D), respectively.

### 2.4 Cord blood genomic DNA isolation and sodium bisulfite modification

Genomic DNA (gDNA) was extracted from whole blood using a blood DNA extraction kit (#MB711400, Lucigen) following the manufacturer’s instructions. DNA integrity was assessed by agarose gel electrophoresis, and DNA purity and concentration were assessed using Nanodrop (Thermo Fisher Scientific, ND1000). Bisulphite conversion of genomic DNA was done using a conversion kit (cat# 59104, Epigentek) according to manufacturer’s protocol. Bisulfite-converted DNA was purified and stored at – 20°C for methylation analysis.

### 2.5 Methylation-specific PCR (MS-PCR)

Leptin gene (*LEP)* sequence was obtained using Ensembl genome browser v.113 (https://asia.ensembl.org/index.html), and the sequence was fed in the MethPrimer 1.0 browser (http://www.urogene.org/methprimer/), where CpG island region was used for primer selection (Figure 1A). Using the Promoter 2.0 server (https://services.healthtech.dtu.dk/services/Promoter-2.0/), the transcription start site (TSS) of *LEP* was located at 800 nucleotides (nt) and denoted as +1. The promoter region generally spans a few hundred base pairs (bp) upstream of TSS. Targeting CpG-rich regions near the TSS for the primer design aids in studying the promoter methylation status of *LEP.* CpG sites situated between -610 and -43 nt relative to the TSS were used for designing the primers. One set of methylated and one set of unmethylated primers were designed in the CpG-rich region of the *LEP* promoter (Figure 1B). *LEP* methylated forward primer was designed at -513 nt, and the methylated reverse primer was designed at -403 nt relative to the position of TSS. *LEP* unmethylated forward and reverse primers were designed at -515 nt and -402 nt, respectively, relative to the TSS.

**Figure 1.**
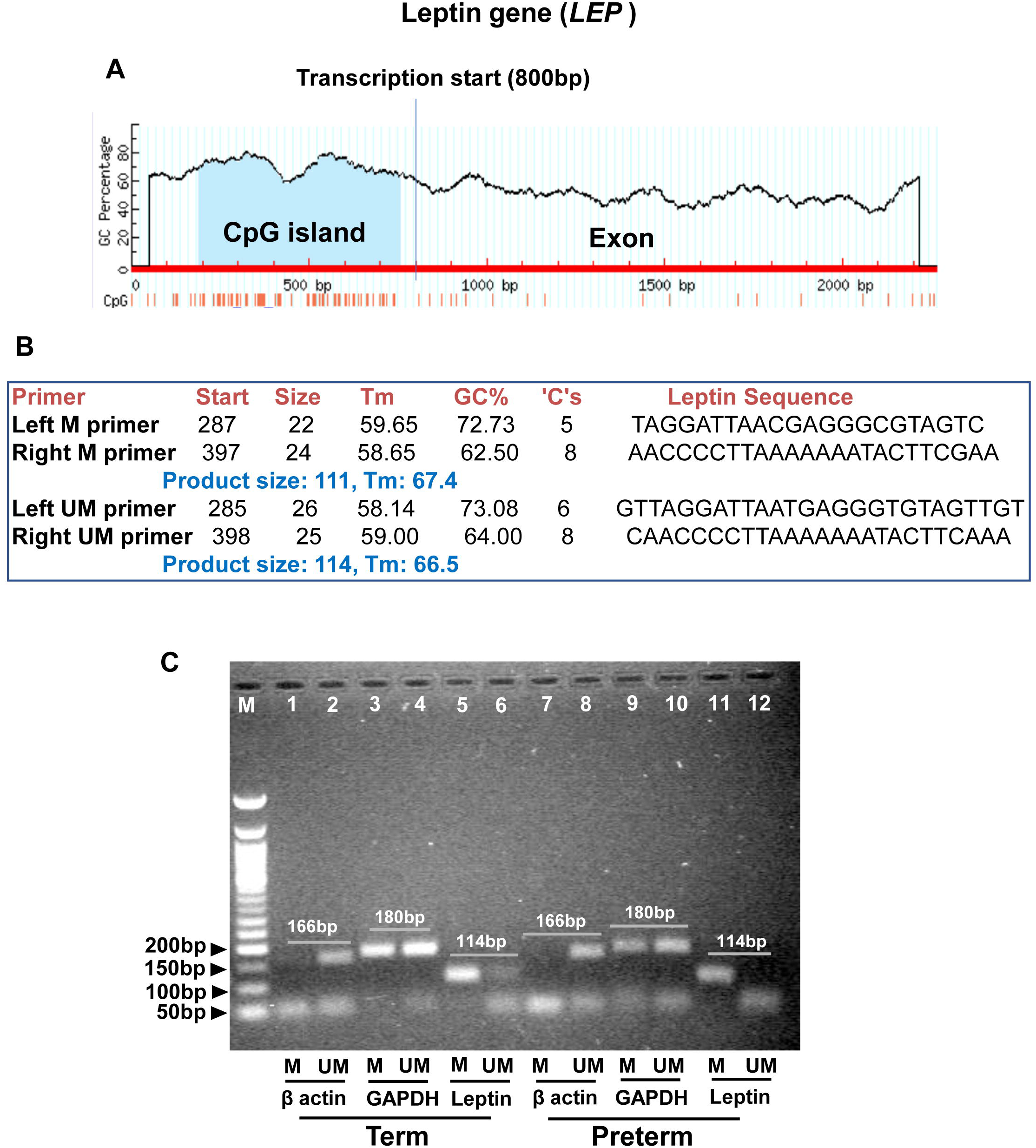
The methylation-specific polymerase chain reaction (MS-PCR) of leptin gene (*LEP*). (A) Design of the leptin methylation-specific primers used in this study. A set of methylated and unmethylated primers were designed by using Meth Primer software, which utilizes the CpG island promoter region of *LEP* to design primers. (B) Primer sequences, amplicon size, Tm, and other features. (C) A representative agarose gel electrophoresis (2%) image indicates the methylation status of the leptin promoter in term and preterm cord blood. Samples were amplified using one methylated (M) and one unmethylated (UM) primer pair. Unlike b-actin, GAPDH was used as the endogenous control as it showed consistent expression in both M and UM wells. M-50 bp ladder; Lane1-6: Term; Lane 7-12: Preterm; Methylation frequency was calculated as the density of the band and expressed as a percentage as indicated in the method. Tm: Melting temperature; GC: Guanine-cytosine

The MS-PCR was performed employing a semi-quantitative-based PCR amplification method where the bisulfite-converted DNA (30 ng) was used as the template. The PCR products were separated by 2% agarose gel (15 cm length). Running a high percentage, long agarose gel, at low voltage for longer duration helps to differentiate methylated and unmethylated DNA products of similar sizes. Human methylated and unmethylated DNA controls (#59695, EpiTect) were used to verify the primer design and efficiency of the bisulfite treatment. PCR amplicons were visualized under UV Transilluminator (GEL DOC 2000 PC, Bio-Rad). The band intensities were measured by Image J software 1.50i (USA) and *LEP* promoter methylation frequencies were calculated (18), as the density of the methylated band/(density of the methylated band + density of the unmethylated band) x100, and the data were expressed as a percentage of methylation frequency.

### 2.6 Cord blood DNA methylation

Global DNA methylation levels were estimated in the cord blood obtained from preterm and term subjects using a methyl flash ELISA kit (#P-1030, Epigentek) as described previously (19).

### 2.7 Total RNA isolation and mRNA expressions by RT-qPCR

Total RNA isolation was performed from the cord blood using the TRI reagent as per the manufacturer’s protocol (Cat# T9424, Sigma). Total RNA was purified using a DNase I kit (#AMPD1, Sigma). The concentration and purity of the total RNA were assessed using Nanodrop (ND1000, Thermo Scientific). The purified total RNA (1000 ng) was converted into cDNA using a kit (#1708891, Bio-Rad), real-time PCR (RT-qPCR) was performed in a 96-well qPCR system (CFX96, Bio-Rad) using TB green pre-mix Ex-Taq II (#RR208, Takara), and KiCqStart SYBR green primers (Table S1, Sigma, Merck). The mRNA expression of the genes was normalized with *PPIB* (endogenous control), and the relative mRNA expression was calculated using the 2^-dCt method.

### 2.8 miRNA target prediction

MicroRNA (miRNA) target prediction of the human *LEP* gene was done using the miRNA target program (20). A total of 2656 miRNAs were targeted to the human leptin gene (NG_007450.1). Subsequently, target network analysis of the obtained hits was done using Cytoscape 3.91.1. The miRNA-target gene network analysis of the top targeted miRNAs for the *LEP* was developed using the Maximal Clique Centrality (MCC) method (21).

### 2.9 Evaluation of oxidative stress by TBARS assay

Cord blood oxidative stress was assessed using Thiobarbituric acid reactive substances (TBARS) assay. Malonaldehyde (MDA) (#108383, Sigma Aldrich) was used as the standard. To serum, 0.3 M Tris-HCl buffer, 20% trichloroacetic acid (#Q28444, Qualigens), and 0.067% thiobarbituric acid (T5500-25G Sigma Aldrich) were added. The samples were spun and the supernatant was used to measure the absorbance at 532 nm using a microplate reader (Biotek, PowerWave XS).

### 2.10 Statistical analysis

Statistical analysis was performed using GraphPad Prism (v 8.0.1). Data with normal distribution was analyzed with a parametric student’s t-test, whereas a non-parametric Mann-Whitney test analyzed data that failed normality. The statistical relationship between the variables was determined by Pearson’s correlation coefficient for normally distributed data and by Spearman correlation for non-normally distributed data. All the values of experimental data were expressed as mean ± standard error of the mean (SEM). P-values < 0.05 were considered statistically significant.

## 3. Results

### 3.1 Preterm deliveries resulted in altered maternal and neonatal parameters

The study included 138 umbilical cord blood samples from neonates; 71 were term, and 67 were preterm. The demographic and clinical parameters of term and preterm mothers and neonates are summarized in Table 1. Gestational age, blood pressure, thyroid-stimulating hormone, and amniotic fluid index were significantly (p<0.05) altered between the two groups. Preterm mothers exhibited substantially lower levels of haemoglobin, red blood cells, and packed cell volume (p<0.05). In preterm mothers, white blood cells and platelets were elevated (p<0.05). Birth weight, APGAR score, head circumference, birth length, and respiratory rate were significantly (p<0.05) reduced in preterm neonates (Table 1). In contrast, heart rate, and cord blood MDA level were significantly (p<0.05) elevated. Since oxidative stress measured by MDA levels is an essential factor in the pathophysiology of preterm birth, preliminary screening for sub-sampling was based on this criterion for further analysis to minimize variability in the dataset.

**Table 1.** Demographic, physiological, and clinical parameters of mothers and neonates.

| Parameter | Term (n=71) | Preterm (n=67) | p-value<sup>\$</sup> |
| --- | --- | --- | --- |
| ***Maternal profile*** |  |  |  |
| Age (years) | 30.03 ± 0.482 | 30.79 ± 0.503 | 0.28<sup>#</sup> |
| Gestational age (weeks) | 38.42 ± 0.093 | 34.89 ± 0.345 | <0.001 |
| Weight (kg) | 72.24 ± 1.435 | 72.59 ± 1.706 | 0.88<sup>#</sup> |
| Systolic pressure (mmHg) | 116.8 ± 1.409 | 124.6 ± 2.095 | 0.007 |
| Diastolic pressure (mmHg) | 72.28 ± 0.914 | 79.06 ± 1.484 | 0.001 |
| Thyroid-stimulating hormone (μU/ml) | 2.202 ± 0.144 | 1.927 ± 0.151 | 0.04 |
| Amniotic fluid index, AFI (cm) | 14.56 ± 0.391 | 12.30 ± 0.491 | <0.001<sup>#</sup> |
| ***Blood profile*** |  |  |  |
| Haemoglobin, Hb (g/dl) | 11.920 ± 0.124 | 11.050 ± 0.197 | <0.001 |
| White blood cells, WBC (10<sup>9</sup>/ml) | 10.210 ± 0.253 | 12.310 ± 1.115 | 0.009 |
| Neutrophils (10<sup>9</sup>/ml) | 7.127 ± 0.223 | 8.367 ± 0.281 | 0.003 |
| Lymphocytes (10<sup>9</sup>/ml) | 1.915 ± 0.029 | 2.170 ± 0.055 | 0.001 |
| Platelets (10<sup>9</sup>/ml) | 224.2 ± 5.538 | 253.2 ± 10.330 | 0.04 |
| Red blood cells, RBC (10<sup>12</sup>/ml) | 4.190 ± 0.048 | 3.945 ± 0.058 | 0.001 |
| Packed cell volume, PCV (%) | 35.88 ± 0.366 | 34.01 ± 0.439 | 0.001 |
| ***Neonatal profile*** |  |  |  |
| Gender | Female (n=34)<br>Male (n=37) | Female (n=31)<br>Male (n=36) | - |
| Birth weight (kg) | 3.00 ± 0.053 | 2.39 ± 0.040 | <0.001 |
| APGAR (1 min) | 8.85 ± 0.088 | 8.19 ± 0.095 | <0.001 |
| APGAR (5 min) | 9.29 ± 0.054 | 8.83 ± 0.089 | <0.001 |
| Head circumference (cm) | 34.35 ± 0.196 | 32.93 ± 0.163 | <0.001 |
| Birth length (cm) | 49.37 ± 0.243 | 47.76 ± 0.274 | <0.001 |
| Heart rate (bpm) | 145.7 ± 1.249 | 150.3 ± 1.153 | 0.009 |
| Respiratory rate (bpm) | 51.73 ± 1.087 | 45.73 ± 1.037 | <0.001 |
| TBARS (nmol MDA/ml) | 1.028 ± 0.041 | 1.277 ± 0.021 | <0.001 |

<sup>\$</sup>The values were expressed as mean ± SEM. (Mann-Whitney test). p<0.05 was considered significant. <sup>#</sup> unpaired t-test. Unpaired t-test and Mann-Whitney were used for normal and non-normal distributed data, respectively. Abbreviation: APGAR: Appearance, Pulse, Grimace, Activity, and Respiration; TBARS: thiobarbituric acid reactive substances; MDA: Malondialdehyde.

### 3.2 Reduced genome-wide methylation and promoter methylation were followed by elevated leptin gene and its receptor expression in preterm cord blood

Global methylation (% 5-methyl cytosine) is significantly lower in preterm neonates [term vs preterm (%): 0.29 ± 0.02 vs 0.18 ± 0.01, p<0.001, Figure 2A]. *LEP*-specific promoter methylation was assessed to comprehend whether the epigenetic dysregulation is associated with leptin in preterm neonates since the latter modulates obesity. Methylation frequency was calculated based on the density of the methylated and unmethylated bands as indicated in the method (Figure 1C). The *LEP*-promoter methylation frequency (%) was significantly reduced in preterm [term vs preterm (%): 72.55 ± 2.833 vs 58.82 ± 2.955, p=0.001, Figure 2B], indicating a likelihood of hypomethylated *LEP*. Moreover, *LEP* methylation was significantly associated (r= 0.594, p=0.02) with global methylation (% 5-mC) in preterm (Figure 2 C-D). In addition, a negative correlation between *LEP* methylation frequency with *LEP* and circulatory leptin in both term and preterm (Figure S1 A-D), indicates that leptin gene methylation might control leptin expression by inhibiting transcription. The relative mRNA expression of both *LEPR* (Figure 2E) and *LEP* (Figure 2F) genes were significantly increased (p=0.04) in preterm, indicating that upregulated mRNA expression was due to the hypomethylation of the *LEP* gene promoter. Although insignificant (p=0.41), the serum leptin showed an increased trend in preterm [term vs preterm (ng/ml): 3.146 ± 0.509 vs 4.095 ± 1.105, Figure 2G]. These findings suggest a potential link between DNA methylation, leptin expression, and altered epigenome signature of preterm neonates.

**Figure 2.**
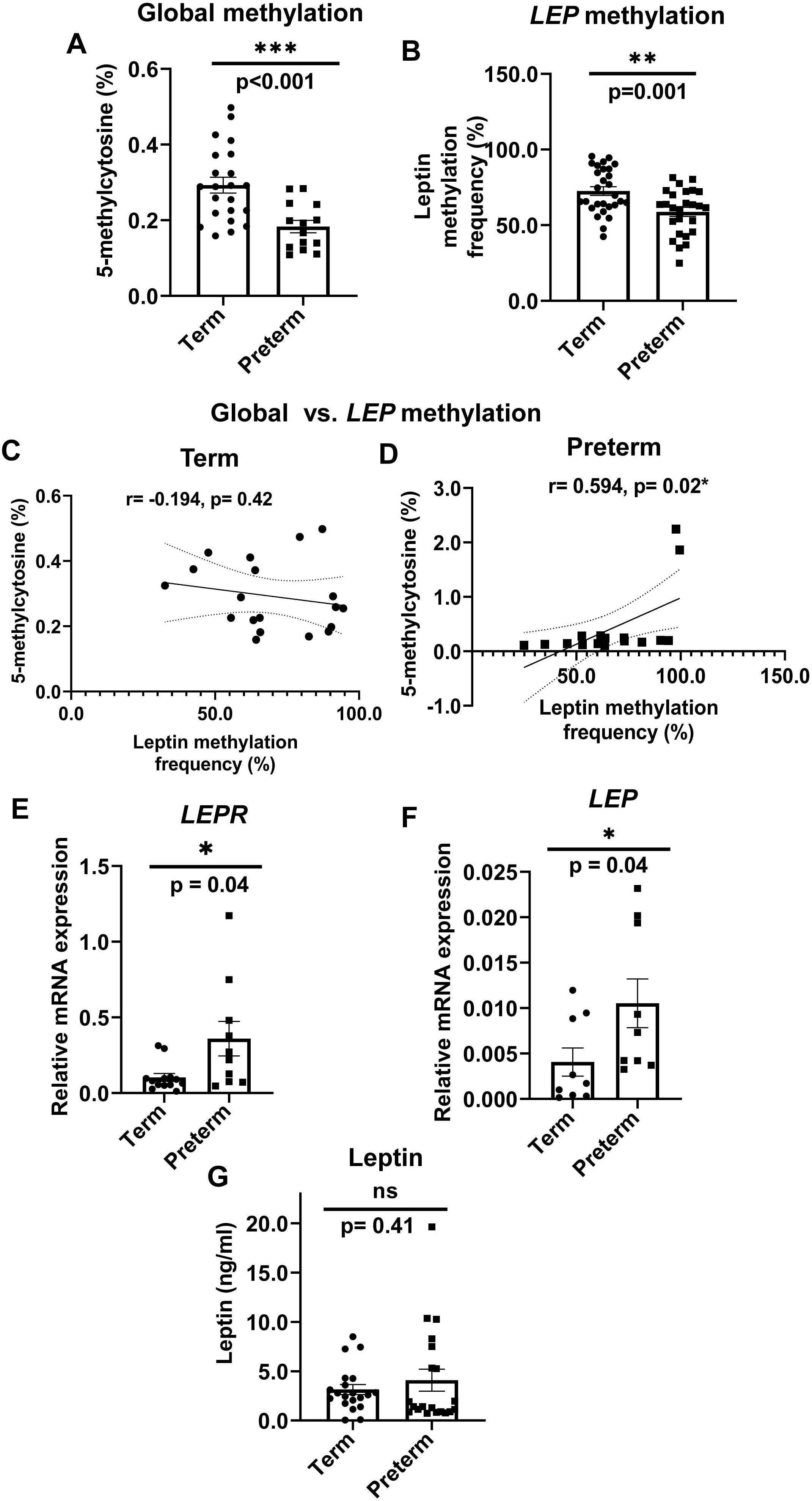
The effects of preterm birth on global and leptin methylation in the neonatal cord blood. (A) Global DNA methylation was assessed from cord blood purified DNA of term (n=22) and preterm (n=14) subjects by measuring the total 5-methylcytosine (5-mC) levels and expressed in percentage. (B) Leptin gene-specific methylation was performed by endpoint PCR using purified cord blood DNA as the template from term (n= 28) and preterm (n= 26) subjects and methylated and unmethylated specific leptin primers. The leptin methylation is expressed as frequencies calculated from the amplicons as indicated in the method. Data are analyzed by an unpaired t-test and expressed as mean ± SEM, where *** represents p<0.001 and **p<0.01. (C-D) Correlation between 5-methylcytosine (%) and leptin methylation frequency (%) in term (n=19) and preterm (n=16) subjects. The statistical relationship between the variables was determined by Pearson’s correlation coefficient (r) and Spearman correlation (p), where * indicates significance if the p-value is less than 0.05. (E-F) Gene expression of leptin receptor (*LEPR)* and leptin (*LEP*) was measured using real-time PCR. Total RNA was purified from term (n=9-13) cord blood and preterm (n=9-10) subjects, converted to cDNA, and run in duplicate using the RT-qPCR method. The gene expression was measured by mRNA levels after normalizing the expression of each mRNA with the expression of endogenous control, *PPIB*. The relative mRNA expression was calculated using the 2^-dCt method and expressed as arbitrary units. (G) Circulatory leptin (term, n=20 and preterm, n=20) was measured in cord blood serum by ELISA. Data was analyzed by the Mann-Whitney test and represented as mean ± SEM. * indicates p < 0.05, and ‘ns’ represents as not significant.

### 3.3. Neonatal birth weight was associated with leptin level but not with its methylation in preterm

The circulatory leptin positively and significantly (r= 0.472, p=0.04) correlated with birth weight in preterm (Figure 3 A-B), implying leptin’s involvement in the initial growth trajectory in preterm neonates. However, birth weight was not dependent on the changes in *LEP* methylation frequency in these neonates (Figure 3 C-D), suggesting that *LEP* promoter methylation might influence neonatal birth weight indirectly by altering circulatory leptin levels. Further, global methylation was inversely correlated (r= -0.455, p=0.04) to term birth weight (Figure 3 E-F), indicating that a lower DNA methylation state of the genome is required in activating genes that might promote birth weight and maintain the growth of the neonate postnatally.

**Figure 3.**
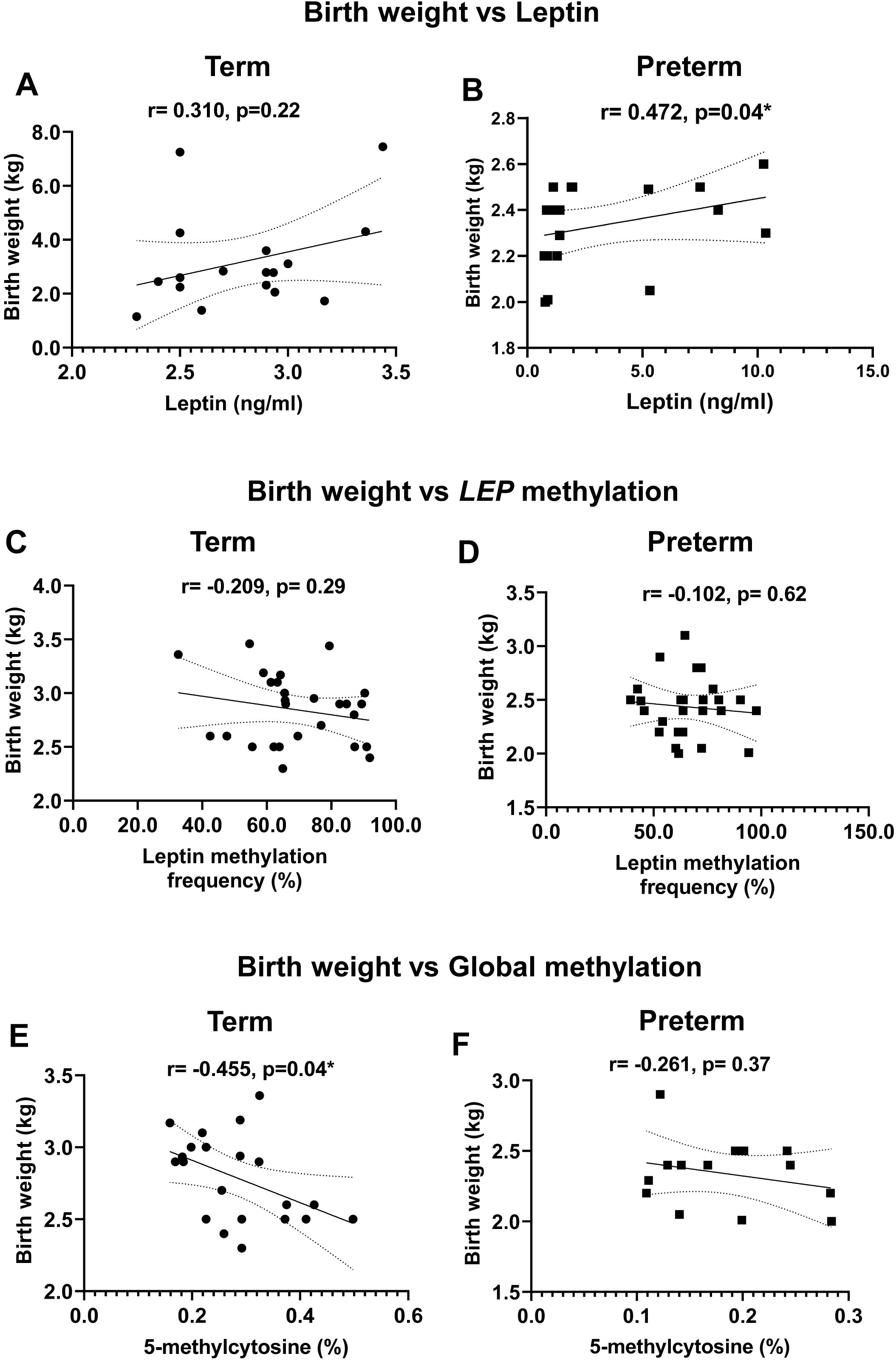
Correlation between neonatal birth weight and global methylation and methylation of leptin gene. Neonatal birth weight was correlated with (A-B) cord blood leptin (term, n=17 and preterm, n=19), (C-D) leptin methylation frequency (term, n=27 and preterm, n=22), and with (E-F) global DNA methylation (term, n= 21 and preterm, n= 14) by Pearson and Spearman correlation coefficients depending on the normality of the data distribution. Each data point depicts an individual subject, and the r-value determines the strength and direction of the association between the variables. The p-value defines statistical significance, and * indicates p < 0.05.

### 3.4 Leptin and its gene methylation collectively stimulated IGF1 action in preterm neonate

Mean IGF1 level was significantly (p=0.01) increased in preterm neonates [term vs preterm (ng/ml): 14.30 ± 1.502 vs. 20.27 ± 1.607, Figure 4A]. IGF1 significantly (r= 0.476, p=0.04) and positively correlated with neonatal birth weight in preterm (Figure 4 B-C), signifying IGF1’s likely role in compensating neonatal catch-up growth. Cord blood IGF1 significantly (r= 0.597, p=0.01) correlated with leptin levels in preterm (Figure 4 D-E). A significant (r= -0.506, p=0.03) inverse association was observed between *LEP* methylation and IGF1 levels in preterm (Figure 4 G-F), suggesting that reduced *LEP* methylation may be associated with promoting IGF1 levels.

**Figure 4.**
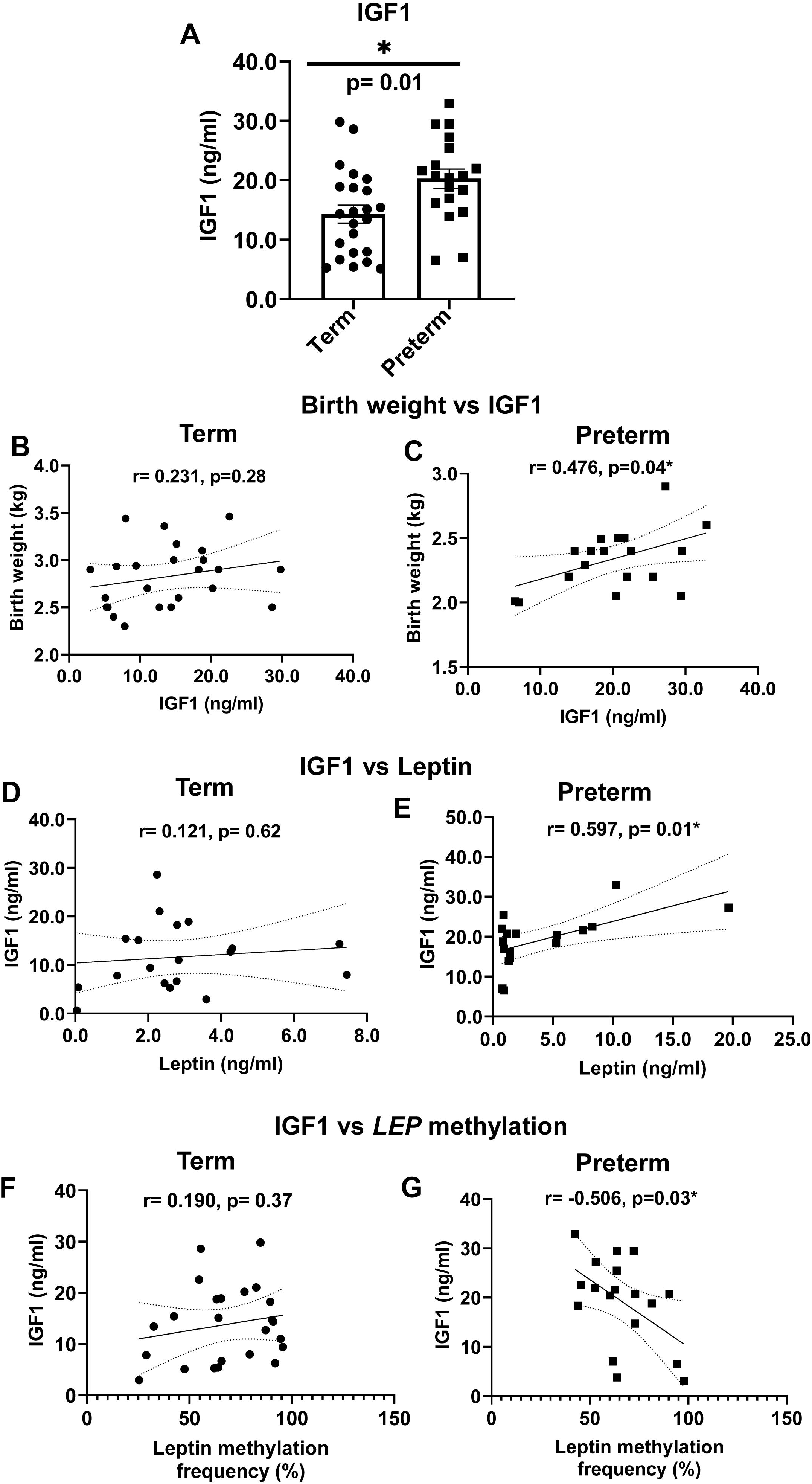
Insulin growth factor (IGF1) levels in cord blood and its association with birth weight, leptin and its methylation in preterm and term subjects. (A) IGF1 level was measured by ELISA and compared between term (n=23) and preterm (n=19) by unpaired t-test. Data was expressed as (ng/ml) mean ± SEM. (B-C) Correlation between IGF1 and neonatal birth weight in (B) term (n=24) and (C) preterm (n=19). Correlation between IGF1 and leptin in (D) term (n=19) and (E) preterm (n=17), and leptin methylation frequency in (F) term (n=24) and (G) preterm (n=18). Pearson’s correlation coefficient determined the statistical association between the variables. The ‘r’ value determines the strength and direction of the association between the variables, and * indicates significance if the p-value is less than 0.05.

### 3.5 Leptin positively correlated with adiponectin expression in regulating the growth of preterm neonates

Adiponectin, leptin, and ghrelin are key adipokines involved in appetite, energy balance and fat reserves that modulate growth hormone release and adiposity. The relative mRNA expression of *ADIPOQ* significantly increased (p<0.001) in preterm cord blood (Figure S2 A) while *GHRL* mRNA expression (Figure S2 B) also showed an increased trend (p>0.05). The correlation between *ADIPOQ* expression and neonatal birth weight showed a counterintuitive inverse trend (p>0.05) in both subjects (Figure S2 C-D). The relative *ADIPOQ* expression was significant (r= 0.716, p=0.01) and positively correlated with *LEP* expression in preterm (Figure S2 E-F).

### 3.6 Leptin positively associates with pro-inflammatory cytokine expression in preterm neonates

The mRNA expression of *IL6* (p=0.03) and *TNF*_α_ (p=0.02) was significantly increased in preterm cord blood (Figure 5 A-B). A correlation analysis was performed between *IL6* and *TNF*_α_ with circulatory leptin, *LEP*, and *LEP* promoter methylation. *TNF*_α_ showed a significant positive correlation (r= 0.644, p=0.02) with circulatory leptin in the preterm (Figure S3 A-B). Unlike term neonate (Figure 5 C, E, and G), *IL6* expression showed a strong positive correlation (r= 0.687, p=0.008) with circulatory leptin, *LEP* (r= 0.763, p=0.009), and an inverse correlation with *LEP* promoter methylation (r= -0.636, p=0.04) in preterm (Figure 5 D, F, and H), indicating that *LEP* expression was associated with the activation of pro-inflammatory cytokines that might disrupt metabolic equilibrium and energy balance.

**Figure 5.**
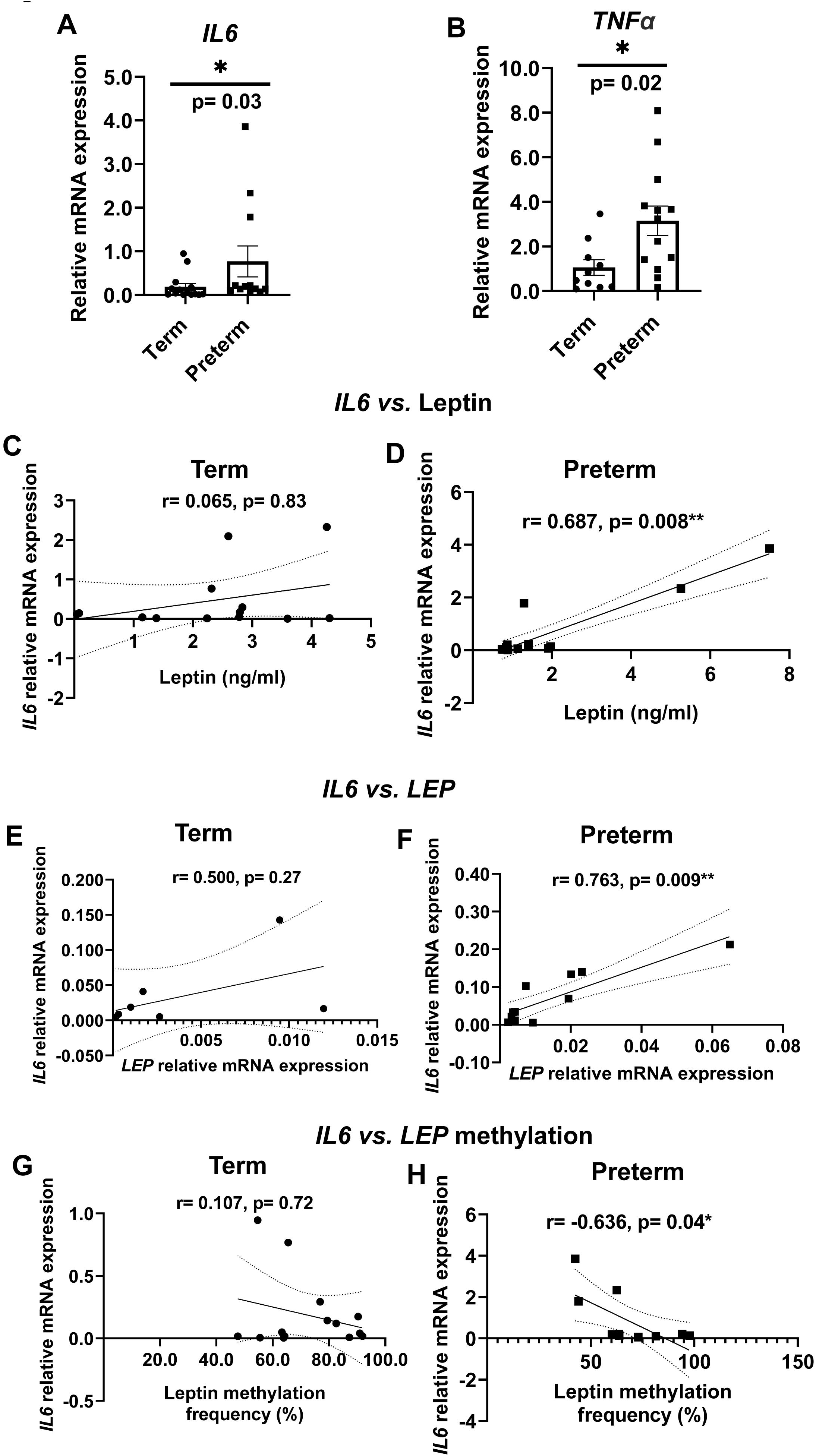
Effect of preterm birth on cord blood inflammatory profile and its association with leptin. (A) *IL6* expression was measured in term (n=14) and preterm (n=12) using RT-qPCR, normalized to endogenous control mRNA levels, and expressed as arbitrary units of relative mRNA expression. The data was analysed using the Mann-Whitney test. (B) *TNF*_α_ mRNA expression was measured in term (n=10) and preterm (n=13) subjects by RT-qPCR, and the data was analysed by unpaired t-test. Cord blood *IL6* was correlated with leptin in (C) term (n=13) and (D) preterm (n=14), with *LEP* in (E) term (n=7), and (F) preterm (n=11), and with leptin methylation frequency in (G) term (n=14) and (H) preterm (n=11) subjects. The statistical association between the variables was determined by the Spearman correlation coefficient. The ‘r’ value specifies the strength and direction of the association between the variables, and * indicates significance if the p-value is less than 0.05, ** indicates the significance if the p-value is less than 0.01.

### 3.7 Neonatal leptin methylation was regulated by gestational age, maternal haemoglobin and systolic blood pressure

Gestational age positively and significantly (r= 0.438, p=0.03) correlated with *LEP* methylation frequency in preterm (Figure S4 A-B), indicating *LEP* methylation could be gestational age-dependent and adjusts the leptin levels through methylations. Moreover, gestational age was positively correlated with leptin (r=0.481, p=0.03) in the term (Figure S4 C), but a weak correlation (p=0.85) in the preterm (Figure S4 D), explaining that maternal adipose reserve contributed due to leptin could be due to increased gestational age. Maternal haemoglobin also positively and significantly (r= 0.448, p=0.05) correlated with *LEP* methylation in preterm (Figure S4 E-F). Maternal (SBP) systolic (r= -0.419, p=0.03) and diastolic (r= -0.351, p=0.08) blood (DBP) pressure was inversely correlated with *LEP* methylation in preterm (Figure S5 A-D), indicating a potential risk of dysregulated blood pressure that *LEP* methylation might transmit to the preterm neonates in their adult life.

### 3.8 MiRNA-dependent regulation of LEP gene and susceptible target pathways associated with obesity

The complete miRNA target for the *LEP* was done using a gap opening penalty of - 9.0, a gap extension penalty of -4.0, and a score threshold of 140. The generated output had 202 hits above the cut-off values (Figure 6A). The top 20 miRNAs based on the total score, position, and stability are presented in Table S2. The specific targeting miRNAs were further identified using the MCC, degree of interaction, closeness, and betweenness (Figure 6B, i-iv). Total target genes by top targeted miRNAs for the *LEP* were considered for analysing their susceptibility to all other human genes. The number of susceptible target genes was highest for has-miR-153-5p (2807). The lowest number of susceptible genes was observed for has-miR-23a-5p (233) (Figure 6C). The top 20 *LEP* gene-targeting miRNAs were identified based on the scores and further analyzed for their respective susceptibility to other gene-targeting pathways, such as body fat distribution, thermogenesis, and adipocyte differentiation (Table 2). Further, the analysis of the top 20 targeted genes by these top 20 miRNAs suggest susceptible genes those possibly targeted by different sets of miRNAs predicted several target pathways including chromatin organization, DNA methylation, transcription regulation including regulation of gene expression, lipid transport and homeostasis, adiponectin-activated signalling pathway, regulation of fat cells, brown fat cell differentiation, energy homeostasis and others (Table S3).

**Figure 6.**
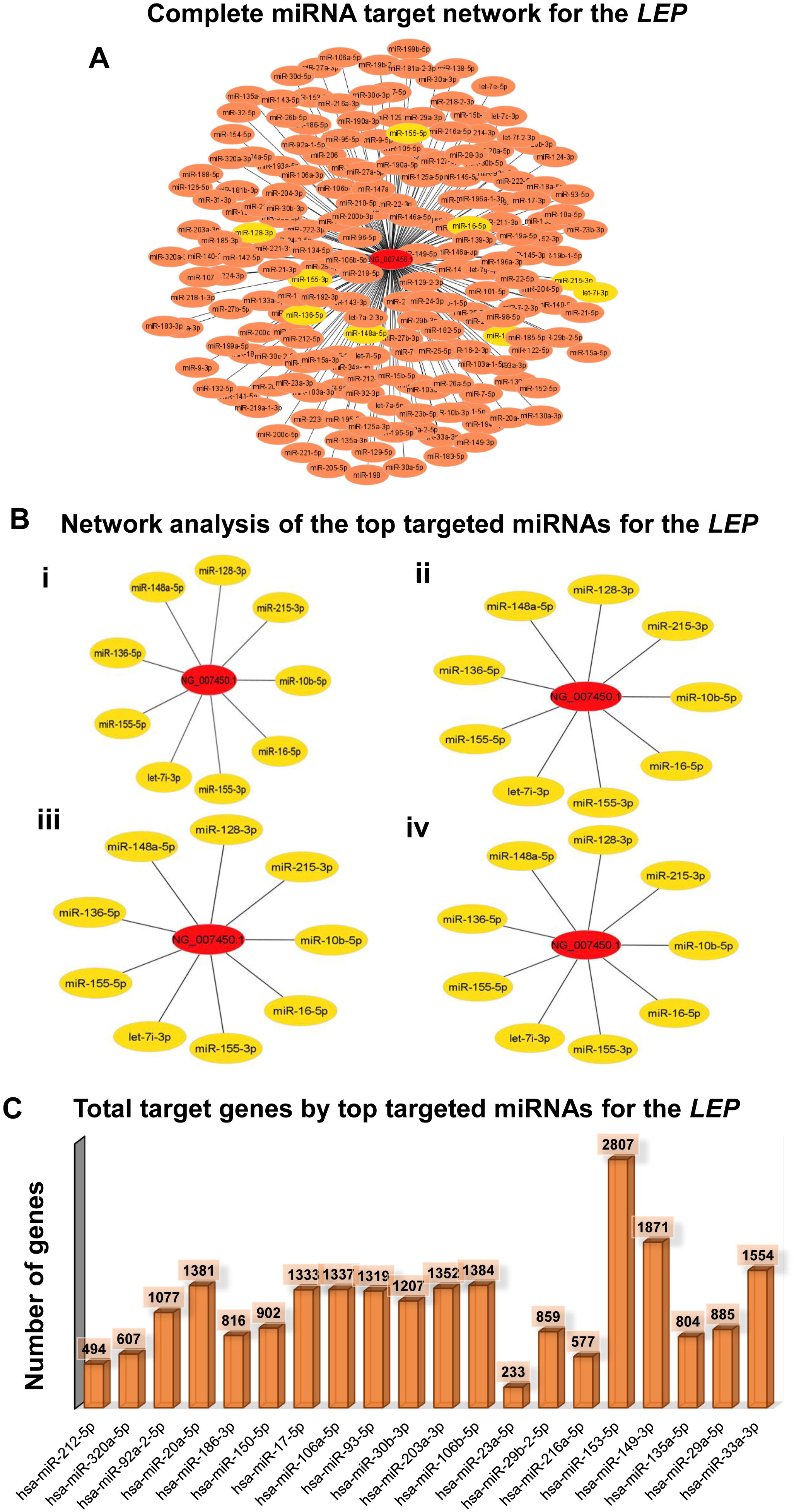
*In silico* analysis of miRNA-dependent regulation of leptin gene and its predictive roles in targeting adiposity and energy metabolism pathways. (A) The complete miRNA target network for the leptin gene (NG007450.1) is represented with an orange circle. In contrast, the yellow circle highlights the top selected miRNAs that are probably highly targeting miRNA as obtained through analysis by the Maximal Clique Centrality method from the entire network. (B) Presentation of the top targeting miRNAs from the overall network analysis as obtained through analysing by (i) maximal Clique Centrality method, (ii) degree of interactions, (iii) closeness, and (iv) betweenness. (C) The number of total target genes by the top 20 miRNAs targeting the leptin gene.

**Table 2.** Top 20 miRNAs shared targets for the top 20 targeting genes.

| **Gene symbol** | **Gene name** | **Predictive target pathways** | **Occurrence** | **Commonly targeted by mRNA** |
| --- | --- | --- | --- | --- |
| *ZNF800* | Zinc finger protein 800 | Endocrine pancreas development, acinar cell differentiation | 6 | miR-20a-5p, miR-17-5p, miR-106a-5p, miR-93-5p, miR-106b-5p, miR-33a-3p |
| *ANKRD52* | Ankyrin repeat domain 52 | Ubiquitous expression in brain, adrenal and other tissues | 5 | miR-20a-5p, miR-17-5p, miR-106a-5p, miR-93-5p, miR-106b-5p |
| *MED12L* | mediator complex subunit 12L | Transcription regulation | 5 | miR-20a-5p, miR-17-5p, miR-106a-5p, miR-93-5p, miR-106b-5p |
| *NPAT* | Nuclear protein, coactivator of histone transcription | Transcription coactivator and corepressor activity | 5 | miR-20a-5p, miR-17-5p, miR-106a-5p, miR-93-5p, miR-106b-5p |
| *TBC1D20* | TBC1 domain family member 20 | Positive regulation of GTPase activity, and lipid droplet organization | 5 | miR-20a-5p, miR-17-5p, miR-106a-5p, miR-93-5p, miR-106b-5p |
| *ZNFX1* | Zinc finger NFX1-type | Activation of innate immunity | 5 | miR-20a-5p, miR-17-5p, miR-106a-5p, miR-93-5p, miR-106b-5p |
| *BRMS1L* | BRMS1 like transcriptional repressor | Regulation of gene expression, cell migration, and growth | 4 | miR-20a-5p, miR-17-5p, miR-106a-5p, miR-93-5p |
| *SAR1B* | Secretion associated Ras related GTPase 1B | Regulation of lipid transport and homeostasis | 4 | miR-20a-5p, miR-17-5p, miR-106a-5p, miR-93-5p |
| *ARID4B* | AT-rich interaction domain 4B | Chromatin organization, DNA methylation, gene expression | 3 | miR-20a-5p, miR-106a-5p, miR-106b-5p |
| *CLOCK* | Clock circadian regulator | Transcription factor activity, circadian clock regulation | 3 | miR-17-5p, miR-106a-5p, miR-93-5p |
| *GPR137C* | G protein-coupled receptor 137C | Positive regulation of TORC1 signaling | 3 | miR-20a-5p, miR-106a-5p, miR-106b-5p |
| *TXNIP* | Thioredoxin interacting protein | Oxidative stress and inflammatory response | 3 | miR-17-5p, miR-106a-5p, miR-93-5p |
| *VLDLR* | Very low-density lipoprotein receptor | VLDL-triglyceride metabolism | 3 | miR-17-5p, miR-106a-5p, miR-93-5p |
| *ADIPOR2* | Adiponectin receptor 2 | Adiponectin-activated signaling pathway | 2 | miR-186-3p, miR-150-5p |
| *DDX6* | DEAD-box helicase 6 | MicroRNA-induced gene silencing | 2 | miR-30b-3p, miR-203a-3p |
| *FOXP4* | Forkhead box P4 | Cell differentiation | 2 | miR-92a-2-5p, miR-149-3p |
| *MAP3K9* | Mitogen-activated protein kinase kinase kinase 9 | Protein phosphorylation, and MAPK cascade | 2 | miR-320a-5p, miR-186-3p |
| *MEX3C* | Mex-3 RNA binding family member C | Regulation of fat cells, and energy homeostasis | 2 | miR-212-5p, miR-203a-3p |
| *NAPEPLD* | N-acyl phosphatidylethanolamine phospholipase D | Phospholipid metabolic process, and regulation of brown fat cell differentiation | 2 | miR-20a-5p, miR-106b-5p |
| *MTCL2* | Microtubule crosslinking factor 2 | Cell cycle, insulin receptor signaling pathway, and negative regulation of gluconeogenesis | 2 | miR-30b-3p, miR-149-3p |

## 4. Discussion

Although this study reports for the first time that *LEP* methylation positively correlated with *IL6* expression in preterm, it is unknown whether promoter methylation of the *LEP* gene would lead to the risk of obesity in the neonate due to chronic inflammation by proinflammatory cytokines. A significant rise in maternal white blood cells (WBC) indicates underlying inflammation that can be passed from mother to fetus for preterm neonates. Further, a significantly elevated *IL6* and *TNF*_α_ expression confirmed the underlying inflammatory state and activated immune system in preterm neonates. The hypomethylated state of global DNA and CpG promoter methylation of *LEP* in preterm neonates observed in our study suggests that they carry altered epigenome fingerprints.

This study demonstrated that preterm neonates exhibited altered adiposity biomarkers, metabolic regulators, and differentiated methylations compared to term. Preterm mothers exhibited higher blood pressure, lower haemoglobin, and increased inflammations, which might have driven them toward preterm delivery. In our study, *LEP* methylation frequency was positively correlated with global methylation in preterm, suggesting that preterm birth carries a signature of altered global epigenome. Leptin was reported to correlate positively with IGF1 only in preterm neonates. Along with a series of other events, like global and *LEP* promoter hypomethylation and increased circulatory leptin, these can contribute to leptin-mediated metabolic deregulation as a result of leptin resistance. The positive correlation between *LEP* expression and proinflammatory mediators (*IL6*, *TNF*_α_) expression can influence each other in promoting the development of low-grade inflammation, as observed in obesity.

Leptin has dual roles as a hormone and a proinflammatory cytokine in regulating adipose metabolism and immune response by modulating inflammation and T-cell activation (22). Leptin levels are typically elevated during infection and inflammation; both are well reported in preterm birth. Higher *LEP*, *LEPR* expression, and circulatory leptin observed in our study can lead to leptin resistance. Dysfunctional *LEPR* and chronic inflammation can lead to leptin resistance (23). Hyperleptinemia and leptin resistance are common factors of obesity. Leptin resistance turns off the functioning of the *LEP*, resulting in elevated appetite and overeating, leading to weight gain and obesity (24). Excess leptin concurrent with *IL6*, *TNF*_α_ expression, and maternal higher WBC observed in this study might promote adipose inflammation and expansion of effector T-cells (25) in preterm neonate resulting in chronic inflammation as observed in obesity.

Promoter methylation of *LEP* typically activates its expression and can trigger leptin-mediated signalling and expression of miRNA. The expression of *LEP* genes regulates multiple factors associated with fat development and controlling white fat cell proliferation. Therefore, the network analyses of the *LEP* gene were performed and obtained top miRNAs that are predicted to target the *LEP* potentially; those are associated with obesity, altered beta cell function (hsa-miR-212-5p) in diabetic conditions (26), key pancreatic gene expression (hsa-miR-92a-2-5p) (27), and others. These miRNAs, such as hsa-miR-212-5p and hsa-miR-92a-2-5p, are potential regulators of the *LEP* expression, and their targeting of the *LEP* may play a significant role in the development of obesity and diabetes. Interestingly, these miRNAs’ most commonly targeted genes also indicated the highly susceptible gene (*ZNF800*) as one of the genes influencing the adiposity-adjusted leptin concentration (28).

Similarly, another top-targeting gene by these miRNAs was *ANKRD52*, which is associated with height, influencing the BMI and obesity estimation (29). Thus, the *LEP* targeting prediction analysis with the available annotated miRNAs provided information on the potential miRNAs that may control the regulation of the *LEP* gene expression. In addition, the study also identified the genes that are probably highly susceptible to these miRNAs and have associated roles related to leptin secretion, obesity, and diabetes.

The dysregulated leptin methylation and global DNA methylation in adipose tissue of subjects with obesity (30) implies that preterm could be at risk for obesity mediated by leptin. *LEP* hypomethylation followed by a higher *LEP* expression might be an adaptive response to lower birth weight (LBW) in preterm neonates observed in this study. Birth weight is an important early indicator for growth trajectory, and LBW preterm neonates pose a risk for dysregulated postnatal adiposity since leptin regulates the fat stores and influences the growth hormone response (31). The increased expression of the *LEP*, *LEPR,* and *IGF1* in preterm cord blood might indicate the altered growth hormone response, which supports the missed growth and developmental phases (32). However, such compensatory growth could elevate the risk of metabolic dysregulation when exposed to overnutrition in later life (33).

Unlike the previous study (34), the present one reported a higher IGF1 in preterm, also associated with leptin. Although IGF1 levels increase with the growth of the infants (35), preterm exhibited higher IGF1 levels, indicating a compensatory growth response. Elevated IGF1 in preterm neonates could result from leptin resistance where the GH/IGF1 axis is dysregulated due to metabolic imbalance (36, 37). Dysfunction or resistance to leptin can lead to elevated fat storage, which stimulates IGF1 production due to increased energy demands. Most small for gestational age children expressed catch-up growth at 18 months, showing a positive correlation with IGF1 (38). Moreover, a positive correlation between *LEP* and *ADIPOQ* expression in this study might signal an additive effect to regulate adiposity in preterm neonates since higher cord blood adiponectin levels are related to central adiposity and paediatric obesity (39, 40).

In our study, we found that preterm mothers exhibit reduced levels of haemoglobin and packed cell volume, which could indicate a dysregulation in neonatal adipose tissue development and altered metabolic programming, similar to what is observed in iron deficiency anaemia (41). We also found that maternal haemoglobin positively correlated with *LEP* methylation, suggesting that the maternal anaemic state might contribute to *LEP* hypomethylation. Another maternal factor, hypertension during pregnancy, resulted in altered cord blood methylation (42). In our study, mothers’ SBP and DBP were negatively correlated with *LEP* methylation frequency, implicating that higher blood pressure of preterm mothers might be one of the contributors to neonatal DNA hypomethylation. It is widely known that LBW in preterm neonates poses several long-term health hazards (43) towards obesity in adult life when accompanied by less physical activity and high-calorie intake (44). Thus, prenatal factors (maternal hypertension, infection), perinatal factors (lower gestational age, lower birth weight), and postnatal factors like formula feeding can further impact the metabolic regulation of preterm neonates. While our study provides meaningful insights, it has a few limitations in the context of small sample size and adopting semi-quantitative MS-PCR. Sample collection was at a single time point, and fetal follow-up couldn’t be achieved in this study.

## 5. Conclusions

Leptin dysregulation observed in preterm cord blood might lead to a chronic inflammatory state that can induce metaflammation and disturb the energy balance regulation due to leptin resistance, resulting in reduced satiety, excess nutrient consumption, and increased total body mass in later life (45). Moreover, the factors contributing to global methylation also impact *LEP* promoter methylation only in the preterm and not in the term.

In the present study, the obtained *LEP* gene-targeting miRNAs also showed affinities for several genes associated with the development of obesity. The observed results confirm the close association of the leptin-miRNA-obesity axis related to fat cell differentiation, fat deposition, and other associations with lipid metabolism that could predict risk in physiological changes of preterm in later obesity. These findings have significant implications for clinical practice, particularly in the early identification and management of metabolic disorders in preterm neonates.

## Data Availability

All data produced in the present study are available upon reasonable request to the authors

## Acknowledgments

Navya Sree Boga was supported by the DST-Inspire Fellowship, Department of Science and Technology, Govt. of India. We appreciate the contribution of KSN Hridayanka during laboratory experiments

## Data availability

Data will be made available on request.

## CRediT authorship contribution statement

Author Contributions: Navya Sree Boga: Investigation, Methodology, Formal analysis, Validation, Software, Writing – original draft, & editing. Amit K Banerjee: In silico data curation, Software, Writing – *in silico* part; Saikanth Varma: Methodology; Archana Molangiri: Methodology; Syeda Farhana: Methodology; Santosh Kumar Banjara: Project administration & planning, Sample procurement, Facilitate collaboration; Nitasha Bagga: Clinical investigation, Sample procurement; Asim K. Duttaroy: Critical comments, Review & Editing; Sanjay Basak: Conceptualization, Supervision, Project administration, Funding acquisition, Formal analysis, Writing – original, review & editing, Finalizing draft;

