## Supplementary figures and images for "Can leptin-specific epigenetic modulation of preterm cord blood predispose obesity?"

### Sup Fig 1

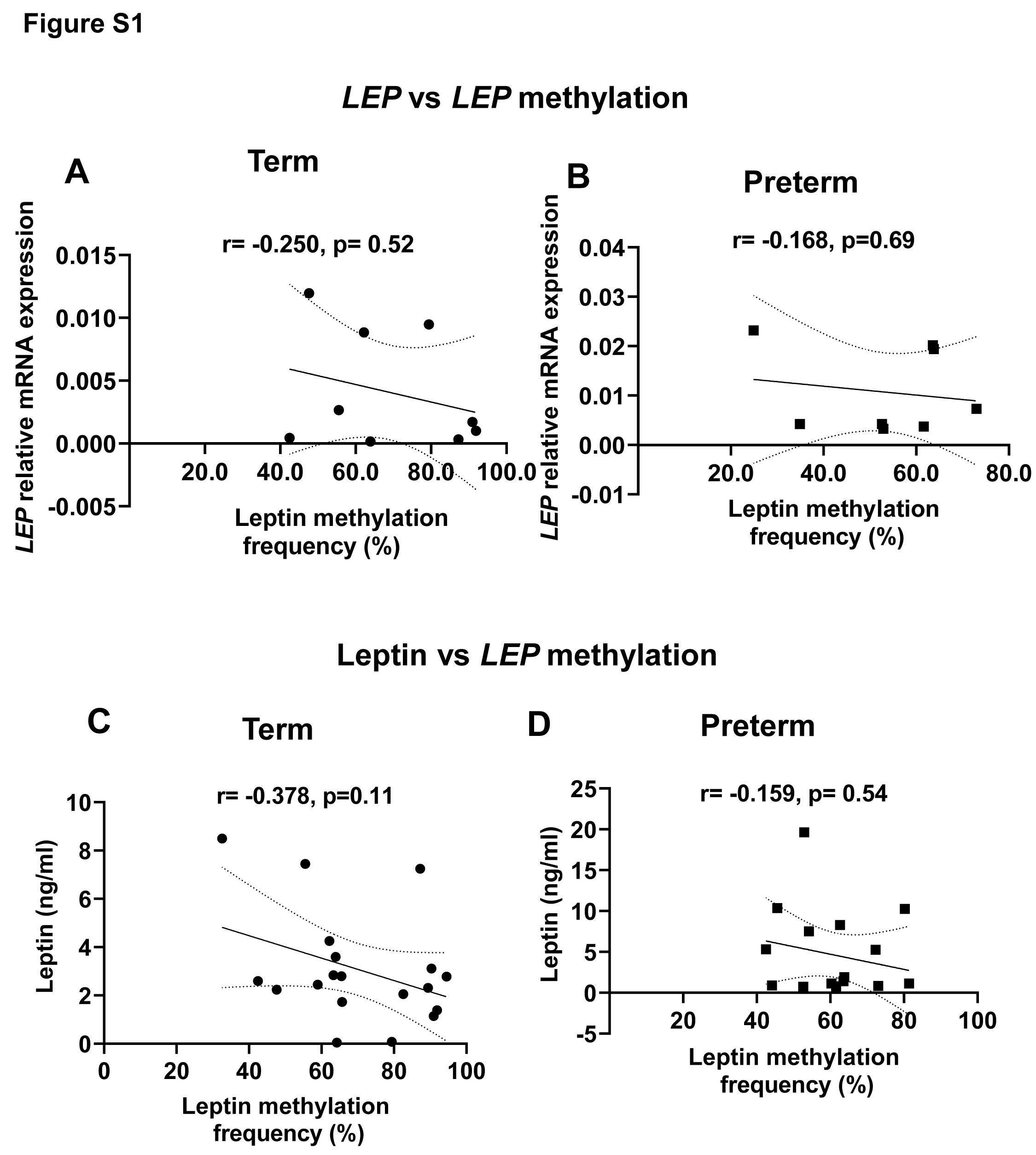

### Sup Fig 2

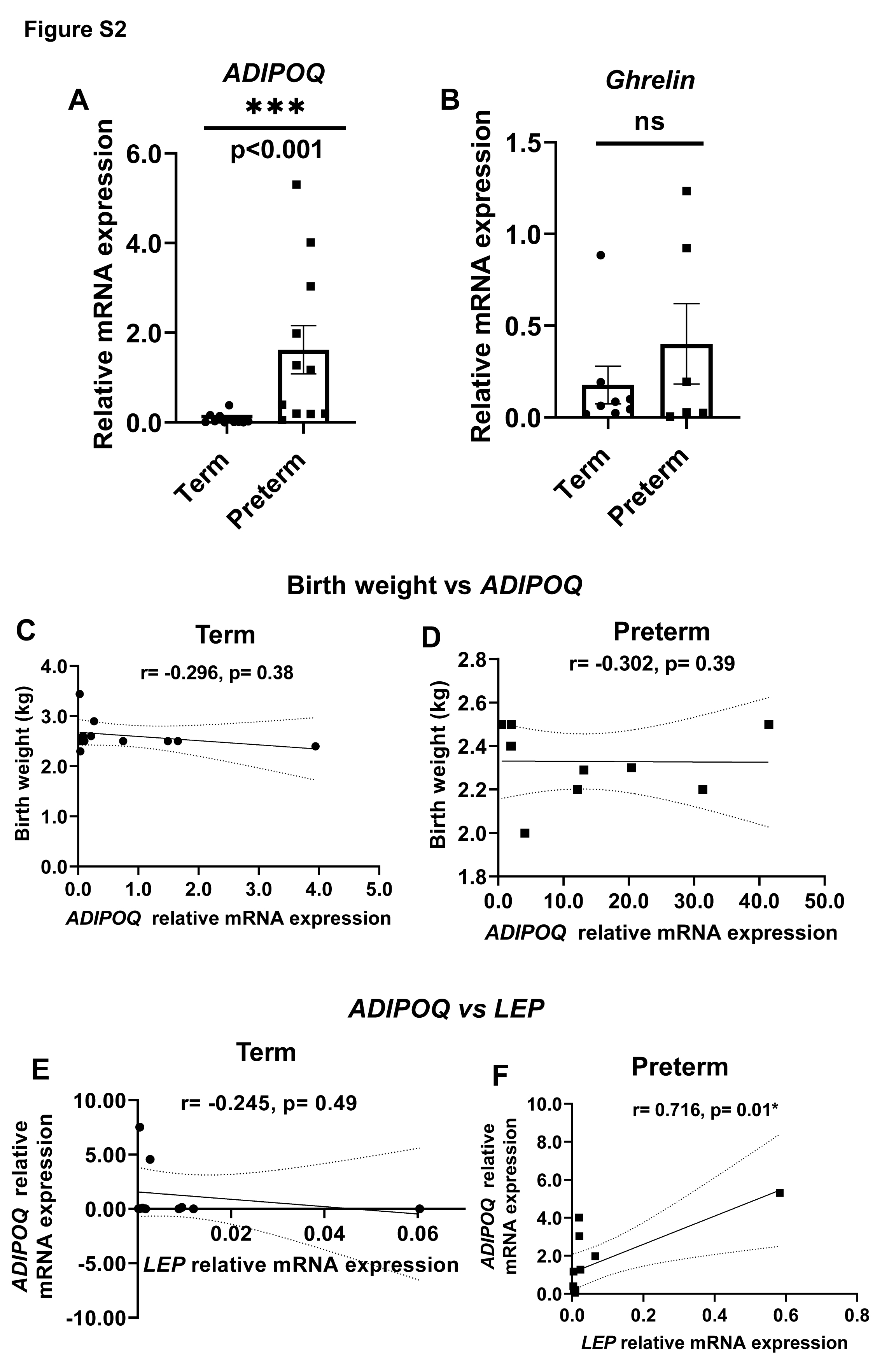

### Sup Fig 3

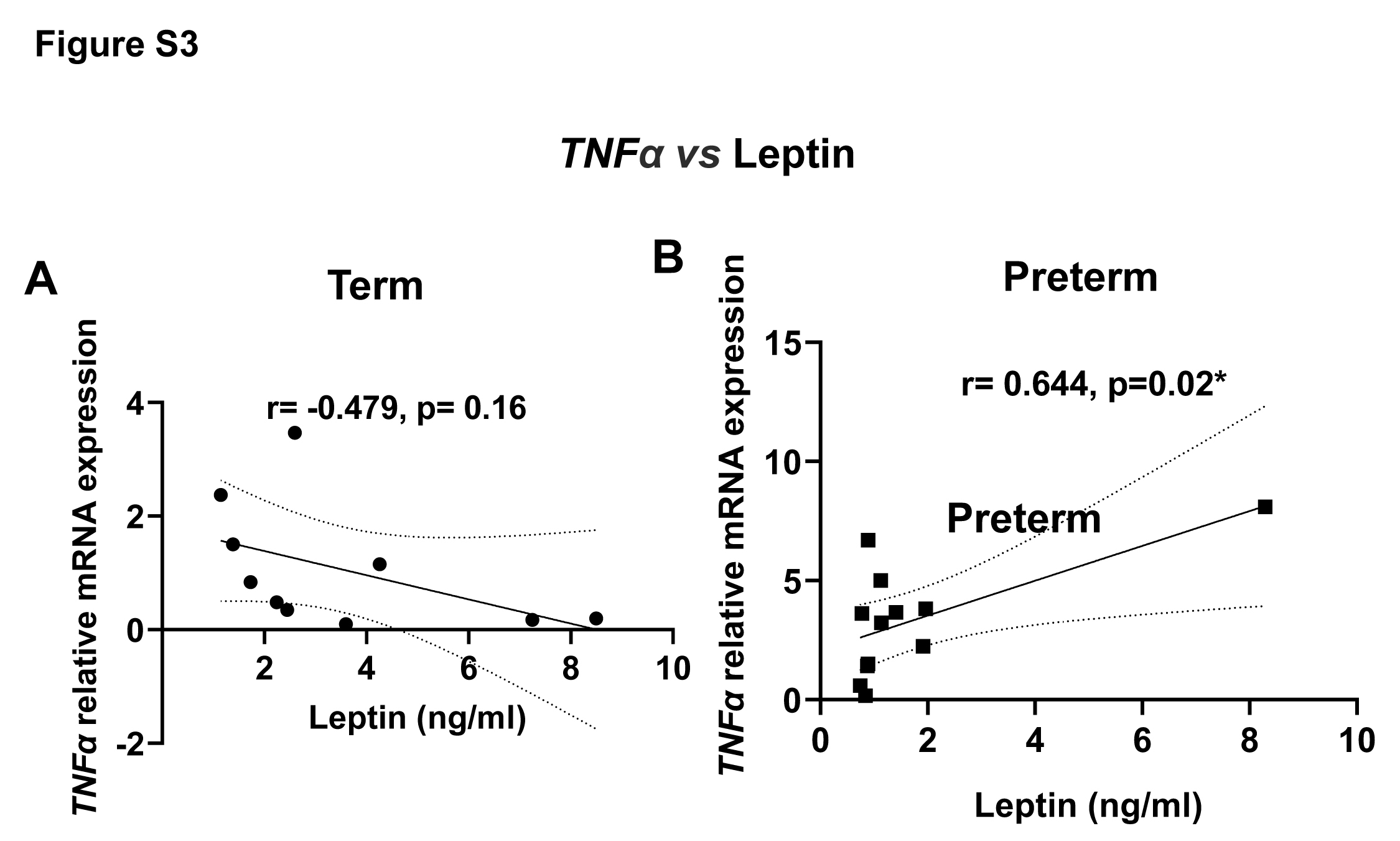

### Sup Fig 4

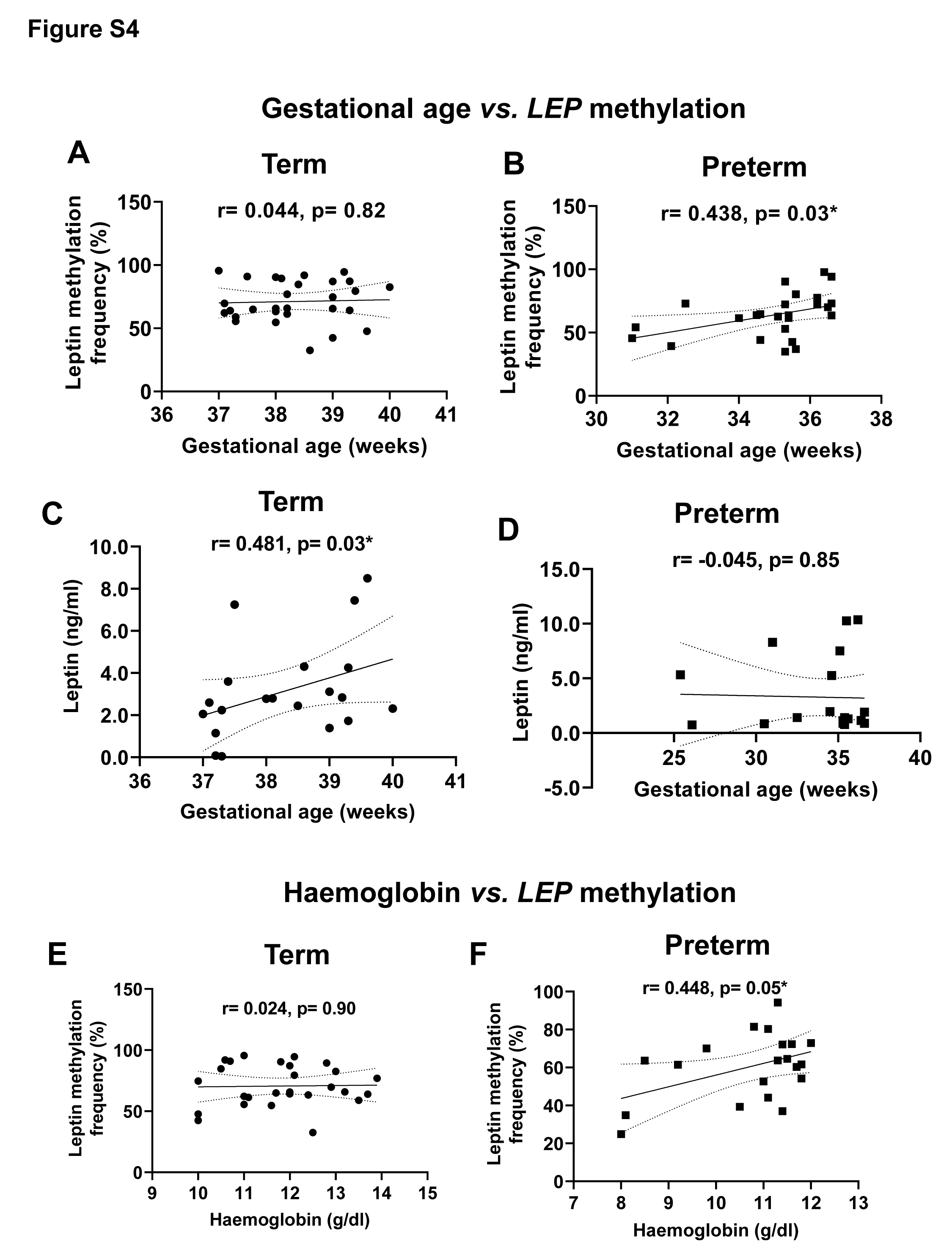

### Sup Fig 5

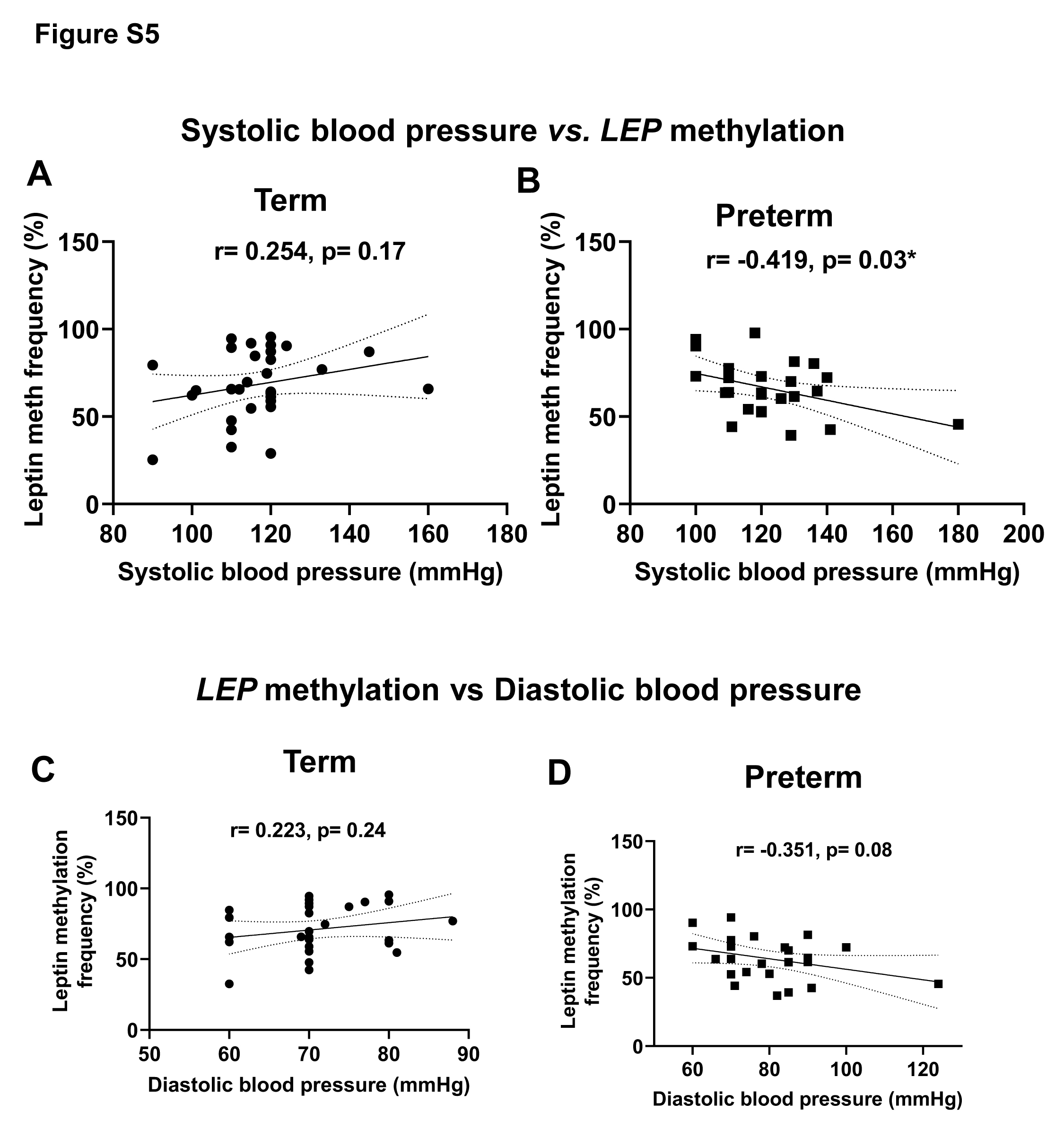
