## Supplementary material for "Can leptin-specific epigenetic modulation of preterm cord blood predispose obesity?": Sup Table 1

**Table S1**. Predesigned SYBR green I human primers and corresponding genes used for the mRNA expression analyses

| **Sl.**  **no.** | **Primer ID** | **Gene symbol** | **Gene ID** | **Gene name** | **Nucleotide sequences (5’-3’)** | **Ref_seqID** |
| --- | --- | --- | --- | --- | --- | --- |
| 1 | H1_LEP | *LEP* | 3952 | Leptin | F 5'- TCAATGACATTTCACACACG-3'  R 5'- TCCATCTTGGATAAGGTCAG -3' | NM_000230 |
| 2 | H1_LEPR | LEPR | 3953 | Leptin receptor | F 5'- ATTTTCAGAAGAGAACGGAC-3'  R 5'- TACTCTATAGACTGTTGGGAAG -3' | NM_001003679 |
| 3 | H1_ADIPOQ | ADIPOQ | 9370 | Adiponectin | F 5'- GGTCTTATTGGTCCTAAGGG-3'  R 5- GTAGAAGATCTTGGTAAAGCG -3' | NM_001177800 |
| 4 | H1_GHRL | GHRL | 51738 | Ghrelin and obestatin prepropeptide | F 5'- GAGAGTCCAGCAGAGAAAG -3'  R 5- GATTCCAACATCAAAGGGG-3' | NM_001134941 |
| 5 | H1-TNFα | TNF | 7124 | Tumor necrosis factor | F 5- CCTCTCTCTAATCAGCCCTCTG-3'  R 5- GAGGACCTGGGAGTAGATGAG-3' | NM_000594 |
| 6 | H1_IL6 | IL6 | 3569 | Interlukin 6 | F 5- ACTCACCTCTTCAGAACGAATTG -3'  R 5- CCATCTTTGGAAGGTTCAGGTTG -3' | NM_000600 |
| 7 | H1_PPIB | PPIB | 5479 | Peptidylprolyl isomerase B | F 5- ACAGGAGAGAAAGGATTTGG -3'  R 5- TGCTTCAGTTTGAAGTTCTC-3' | NM_000942 |
