## Supplementary material for "Can leptin-specific epigenetic modulation of preterm cord blood predispose obesity?": Sup Table 2

**Table S2.** Obtained top 20 predicted human miRNAs targeting the human leptin gene (NG007450.1)

| **miRNA** | **Total Score** | **Total Energy** | **Maximum Score** | **Maximum Energy** | **Strand** | **miRNA length (bp)** | **Gene length (bp)** | **Positions** | **Positions** | **Positions** | **Positions** |
| --- | --- | --- | --- | --- | --- | --- | --- | --- | --- | --- | --- |
| hsa-miR-212-5p | 2723 | -392.01 | 171 | -26.27 | 144 | 23 | 23351 | 2270 | 7718 | 8700 | 11900 |
| hsa-miR-320a-5p | 2631 | -404.75 | 156 | -33.44 | 260 | 22 | 23351 | 22932 | 8622 | 16000 | 18509 |
| hsa-miR-92a-2-5p | 2059 | -294.37 | 160 | -27.11 | 61 | 22 | 23351 | 18223 | 6724 | 21975 | 7783 |
| hsa-miR-20a-5p | 1969 | -245.34 | 169 | -22.68 | 28 | 23 | 23351 | 11890 | 21030 | 16189 | 21900 |
| hsa-miR-186-3p | 1894 | -237.34 | 160 | -20.82 | 249 | 22 | 23351 | 5956 | 5842 | 7709 | 14254 |
| hsa-miR-150-5p | 1854 | -243.85 | 158 | -35.77 | 241 | 22 | 23351 | 17644 | 13270 | 7847 | 4763 |
| hsa-miR-17-5p | 1840 | -249.43 | 169 | -24.79 | 19 | 23 | 23351 | 11890 | 21030 | 16189 | 7341 |
| hsa-miR-106a-5p | 1840 | -212.16 | 169 | -24.33 | 84 | 23 | 23351 | 11890 | 21030 | 16189 | 7341 |
| hsa-miR-93-5p | 1830 | -263.69 | 169 | -28.17 | 62 | 23 | 23351 | 11890 | 21030 | 16189 | 21900 |
| hsa-miR-30b-3p | 1791 | -233.78 | 157 | -27.4 | 182 | 22 | 23351 | 1793 | 12777 | 3731 | 9021 |
| hsa-miR-203a-3p | 1776 | -167.68 | 155 | -21.18 | 135 | 22 | 23351 | 6999 | 5901 | 10780 | 15858 |
| hsa-miR-106b-5p | 1694 | -189.82 | 167 | -20.77 | 270 | 21 | 23351 | 11892 | 21032 | 16191 | 21902 |
| hsa-miR-23a-5p | 1597 | -309.3 | 151 | -38.04 | 34 | 22 | 23351 | 18924 | 13038 | 9033 | 11062 |
| hsa-miR-29b-2-5p | 1496 | -186.34 | 160 | -23.83 | 78 | 22 | 23351 | 17820 | 18646 | 11822 | 21098 |
| hsa-miR-216a-5p | 1467 | -169.87 | 167 | -21.11 | 151 | 22 | 23351 | 16066 | 10143 | 21852 | 3470 |
| hsa-miR-153-5p | 1462 | -99.26 | 155 | -13.2 | 219 | 22 | 23351 | 11849 | 16284 | 338 | 1857 |
| hsa-miR-149-3p | 1462 | -261.49 | 160 | -31.46 | 240 | 21 | 23351 | 11226 | 12794 | 9215 | 4810 |
| hsa-miR-135a-5p | 1459 | -155.47 | 163 | -22.92 | 197 | 23 | 23351 | 19581 | 8326 | 1019 | 12481 |
| hsa-miR-29a-5p | 1450 | -121.27 | 161 | -19.51 | 49 | 22 | 23351 | 2738 | 1858 | 17458 | 12159 |
| hsa-miR-33a-3p | 1448 | -97.39 | 150 | -17.92 | 58 | 22 | 23351 | 14335 | 11143 | 13933 | 1666 |
