## Supplementary material for "Can leptin-specific epigenetic modulation of preterm cord blood predispose obesity?": Sup Table 3

**Table S3.** Top 20 miRNAs shared targets for the top 20 targeting genes and associated pathway

| **Genes** | **Associated Human pathways (BioCyc GO Terms)** | **Occurrence** | **Commonly targeted by mRNA** |
| --- | --- | --- | --- |
| *ZNF800* | Regulation of DNA-templated transcription, endocrine pancreas development, acinar cell differentiation | 6 | miR-20a-5p, miR-17-5p, miR-106a-5p, miR-93-5p, miR-106b-5p, miR-33a-3p |
| *ANKRD52* | Regulation of barbed-end actin filament capping | 5 | miR-20a-5p, miR-17-5p, miR-106a-5p, miR-93-5p, miR-106b-5p |
| *DYNC1LI2* | Microtubule cytoskeleton organization, microtubule-based process, centrosome localization, cellular response to nerve growth factor stimulus | 5 | miR-20a-5p, miR-17-5p, miR-106a-5p, miR-93-5p, miR-106b-5p |
| *ENPP5* | Cell communication, nucleotide metabolic process | 5 | miR-20a-5p, miR-17-5p, miR-106a-5p, miR-93-5p, miR-106b-5p |
| *FYCO1* | Plus-end-directed vesicle transport along microtubule, positive regulation of autophagosome maturation, 'de novo' protein folding, chaperone-mediated protein folding | 5 | miR-20a-5p, miR-17-5p, miR-106a-5p, miR-93-5p, miR-106b-5p |
| *KCNB1* | Potassium ion transmembrane transport, positive regulation of protein targeting to membrane, monoatomic ion transport, potassium ion transport, exocytosis vesicle docking involved in exocytosis, glutamate receptor signaling pathway positive regulation of norepinephrine secretion, cellular response to nutrient levels positive regulation of catecholamine secretion, monoatomic ion transmembrane transport glucose homeostasis, clustering of voltage-gated potassium channels, positive regulation of calcium ion-dependent exocytosis, negative regulation of insulin secretion, response to axon injury, protein homooligomerization, transmembrane transport, cellular response to calcium ion, cellular response to glucose stimulus protein localization to plasma membrane, potassium ion export across the plasma membrane regulation of action potential, positive regulation of long-term synaptic depression response to L-glutamate, regulation of motor neuron apoptotic process | 5 | miR-320a-5p, miR-17-5p, miR-106a-5p, miR-93-5p, miR-135a-5p |
| *MED12L* | DNA-templated transcription, regulation of transcription by RNA polymerase II, positive regulation of DNA-templated transcription, positive regulation of transcription by RNA polymerase II | 5 | miR-20a-5p, miR-17-5p, miR-106a-5p, miR-93-5p, miR-106b-5p |
| *NPAS2* | DNA damage response, positive regulation of DNA repair, positive regulation of DNA-templated transcription, positive regulation of transcription by RNA polymerase II, response to redox state, regulation of DNA-templated transcription, regulation of DNA-templated transcription, central nervous system development, response to xenobiotic stimulus, circadian regulation of gene expression, rhythmic process, positive regulation of behavioral fear response | 5 | miR-20a-5p, miR-17-5p, miR-106a-5p, miR-93-5p, miR-106b-5p |
| *NPAT* | Regulation of gene expression, cell cycle G1/S phase transition, positive regulation of DNA-templated transcription, positive regulation of transcription by RNA polymerase II, in utero embryonic development, DNA-templated transcription, cell cycle, negative regulation of DNA-templated transcription | 5 | miR-20a-5p, miR-17-5p, miR-106a-5p, miR-93-5p, miR-106b-5p |
| *TBC1D20* | Endoplasmic reticulum to Golgi vesicle-mediated transport, Golgi organization, positive regulation of GTPase activity, positive regulation by host of viral genome replication, positive regulation by virus of viral protein levels in host cell, COPII-coated vesicle cargo loading, regulation of cilium assembly, positive regulation of ER to Golgi vesicle-mediated transport, acrosome assembly, lens development in camera-type eye, spermatogenesis, male gonad development, viral process, vesicle-mediated transport, lipid droplet organization, lens fiber cell morphogenesis, seminiferous tubule development | 5 | miR-20a-5p, miR-17-5p, miR-106a-5p, miR-93-5p, miR-106b-5p |
| *ZNFX1* | Defense response to bacterium, defense response to virus, activation of innate immune response, immune system process, regulation of DNA-templated transcription, regulatory ncRNA-mediated heterochromatin formation, pericentric heterochromatin formation, negative regulation of viral genome replication | 5 | miR-20a-5p, miR-17-5p, miR-106a-5p, miR-93-5p, miR-106b-5p |
| *BRMS1L* | Negative regulation of transcription by RNA polymerase II, regulation of gene expression, negative regulation of cell migration, negative regulation of transforming growth factor beta receptor signaling pathway, regulation of growth, negative regulation of stem cell population maintenance, positive regulation of stem cell population maintenance | 4 | miR-20a-5p, miR-17-5p, miR-106a-5p, miR-93-5p |
| *SAR1B* | endoplasmic reticulum to Golgi vesicle-mediated transport, regulation of lipid transport, lipoprotein transport, COPII vesicle coating, lipid homeostasis, COPII-coated vesicle cargo loading,  regulation of TORC1 signaling, negative regulation of TORC1 signaling, cellular response to leucine starvation, regulation of COPII vesicle coating, regulation of COPII vesicle coating, intracellular protein transport, intracellular protein transport, small GTPase-mediated signal transduction, protein transport, vesicle organization, vesicle-mediated transport, membrane organization, positive regulation of protein exit from endoplasmic reticulum, lipid export from cell | 4 | miR-20a-5p, miR-17-5p, miR-106a-5p, miR-93-5p |
| *ARID4B* | Negative regulation of transcription by RNA polymerase II, DNA methylation, chromatin organization, chromatin organization, regulation of DNA-templated transcription, regulation of transcription by RNA polymerase II, spermatogenesis, regulation of gene expression, negative regulation of cell migration, negative regulation of transforming growth factor beta receptor signaling pathway, positive regulation of transcription by RNA polymerase II, regulation of nitrogen compound metabolic process, genomic imprinting, regulation of primary metabolic process,  establishment of Sertoli cell barrier, negative regulation of stem cell population maintenance. | 3 | miR-20a-5p, miR-106a-5p, miR-106b-5p |
| *CLOCK* | Entrainment of circadian clock, entrainment of circadian clock by photoperiod, circadian regulation of translation | 3 | miR-17-5p, miR-106a-5p, miR-93-5p |
| *GPR137C* | Positive regulation of TORC1 signaling | 3 | miR-20a-5p, miR-106a-5p, miR-106b-5p |
| *GUCY1A2* | cGMP biosynthetic process, signal transduction, nitric oxide mediated signal transduction, cyclic nucleotide biosynthetic process, positive regulation of nitric oxide mediated signal transduction, intracellular signal transduction, response to oxygen levels | 3 | miR-203a-3p, miR-29b-2-5p, miR-33a-3p |
| *ITGB8* | vasculogenesis, cell-matrix adhesion, integrin-mediated signaling pathway, integrin-mediated signaling pathway, positive and negative regulation of gene expression, positive regulation of angiogenesis, cartilage development, regulation of transforming growth factor beta activation, ganglioside metabolic process, immune response, transforming growth factor beta receptor signaling pathway, multicellular organism development, response to virus, cell migration, cell adhesion mediated by integrin, hard palate development, placenta blood vessel development Langerhans cell differentiation, cell-cell adhesion | 3 | miR-20a-5p, miR-17-5p, miR-106a-5p |
| *RUFY2* | Regulation of endocytosis | 3 | miR-17-5p, miR-106a-5p, miR-93-5p |
| *TXNIP* | Response to oxidative stress, keratinocyte differentiation, negative regulation of cell division, cellular response to tumor cell, negative regulation of transcription by RNA polymerase II protein import into nucleus, inflammatory response, cell cycle, response to xenobiotic stimulus, response to mechanical stimulus, response to glucose, protein transport, response to estradiol, response to progesterone, regulation of cell population proliferation, response to hydrogen peroxide, positive regulation of apoptotic process, negative regulation of catalytic activity, platelet-derived growth factor receptor signaling pathway, response to calcium ion, cellular response to oxidised low-density lipoprotein particle stimulus | 3 | miR-17-5p, miR-106a-5p, miR-93-5p |
| *VLDLR* | Regulation of very-low-density lipoprotein particle remodeling, Positive and negative regulation of very-low-density lipoprotein particle remodeling | 3 | miR-17-5p, miR-106a-5p, miR-93-5p |
| *ADIPOR2* | Adiponectin-activated signaling pathway, adiponectin-activated signaling pathway, positive regulation of protein phosphorylation, lipid metabolic process, fatty acid metabolic process, heart development, female pregnancy, response to nutrient, response to xenobiotic stimulus, response to bacterium, response to fructose, hormone-mediated signaling pathway, negative regulation of gene expression, response to amine, fatty acid oxidation, negative regulation of cell growth, response to nutrient levels, response to lipopolysaccharide, glucose homeostasis, glucose homeostasis, response to ethanol, positive regulation of glucose import, vascular wound healing, vascular wound healing, negative regulation of hepatic stellate cell migration, cellular response to fatty acid, positive regulation of cold-induced thermogenesis, positive regulation of cold-induced thermogenesis | 2 | miR-186-3p, miR-150-5p |
| *AR* | No hits | 2 | miR-92a-2-5p, miR-149-3p |
| *ARHGAP12* | Morphogenesis of an epithelial sheet, phagocytosis, engulfment actin filament organization, negative regulation of small GTPase mediated signal transduction signal transduction, regulation of GTPase activity, positive regulation of GTPase activity | 2 | miR-17-5p, miR-93-5p |
| *B3GLCT* | Carbohydrate metabolic process, fucose metabolic process, protein glycosylation | 2 | miR-135a-5p, miR-29a-5p |
| *CADM2* | Cell adhesion, homophilic cell adhesion via plasma membrane adhesion molecules, homophilic cell adhesion via plasma membrane adhesion molecules, heterophilic cell-cell adhesion via plasma membrane cell adhesion molecules, brain development, cell recognition | 2 | miR-216a-5p, miR-33a-3p |
| *DDX6* | Negative regulation of translation, viral RNA genome packaging, P-body assembly, miRNA-mediated gene silencing by inhibition of translation, regulation of translation, spermatogenesis, RNA secondary structure unwinding, stem cell population maintenance, neuron differentiation, stress granule assembly, stress granule assembly, negative regulation of neuron differentiation, spermatid differentiation | 2 | miR-30b-3p, miR-203a-3p |
| *FOXP4* | Negative regulation of transcription by RNA polymerase II, regulation of DNA-templated transcription, regulation of transcription by RNA polymerase II, anatomical structure morphogenesis, cell differentiation | 2 | miR-92a-2-5p, miR-149-3p |
| *HOOK3* | Endosome organization, lysosome organization, endosome to lysosome transport, cytoplasmic microtubule organization, early endosome to late endosome transport, Golgi localization, protein localization to perinuclear region of cytoplasm, Golgi organization, protein transport, interkinetic nuclear migration, interkinetic nuclear migration, cytoskeleton-dependent intracellular transport, microtubule anchoring at centrosome, negative regulation of neurogenesis, protein localization to centrosome, neuronal stem cell population maintenance | 2 | miR-30b-3p, miR-33a-3p |
| *MAP3K9* | Protein phosphorylation, protein autophosphorylation, MAPK cascade, apoptotic process, signal transduction, cell death, positive regulation of apoptotic process | 2 | miR-320a-5p, miR-186-3p |
| *MEX3C* | Chondrocyte hypertrophy, protein ubiquitination, regulation of fat cell, differentiation, energy homeostasis | 2 | miR-212-5p, miR-203a-3p |
| *NACC1* | Positive regulation of cell population proliferation, negative regulation of DNA-templated transcription, protein homooligomerization, negative regulation of transcription by RNA polymerase II, regulation of transcription by RNA polymerase II | 2 | miR-92a-2-5p, miR-149-3p |
| *NAPEPLD* | N-acylphosphatidylethanolamine metabolic process, temperature homeostasis, lipid metabolic process, phospholipid metabolic process, phospholipid catabolic process, lipid catabolic process, response to isolation stress, host-mediated regulation of intestinal microbiota composition, host-mediated regulation of intestinal microbiota composition, positive regulation of inflammatory response, N-acylethanolamine metabolic process, N-acylethanolamine metabolic process, positive regulation of brown fat cell differentiation, positive regulation of brown fat cell differentiation, negative regulation of eating behavior | 2 | miR-20a-5p, miR-106b-5p |
| *NFIX* | Negative regulation of transcription by RNA polymerase II, DNA replication, regulation of DNA-templated transcription, regulation of transcription by RNA polymerase II, transcription by RNA polymerase II, positive regulation of DNA-templated transcription | 2 | miR-92a-2-5p, miR-149-3p |
| *PAPOLG* | DNA-templated transcription, mRNA polyadenylation, mRNA processing, RNA 3'-end processing | 2 | miR-30b-3p, miR-149-3p |
| *PHF20L1* | Negative regulation of proteasomal ubiquitin-dependent protein catabolic process, negative regulation of protein catabolic process, regulation of DNA-templated transcription, regulation of transcription by RNA polymerase II | 2 | miR-212-5p, miR-216a-5p |
| *PKD2* | Branching involved in ureteric bud morphogenesis, liver development, heart looping, calcium ion transport, cell surface receptor signaling pathway, heart development, Wnt signaling pathway, spinal cord development, neural tube development, positive regulation of inositol 1,4,5-trisphosphate-sensitive calcium-release channel activity, metanephric part of ureteric bud development, sodium ion transmembrane transport, aorta development, regulation of cell population proliferation, cytoplasmic sequestering of transcription factor, cilium organization, positive regulation of nitric oxide biosynthetic process, positive regulation of cyclin-dependent protein serine/threonine kinase activity, positive regulation of transcription by RNA polymerase II, release of sequestered calcium ion into cytosol, protein homo and hetero tetramerization, regulation of cell cycle, renal artery morphogenesis, calcium ion transmembrane transport, response to cAMP, cellular response to hydrostatic pressure, cellular response to osmotic stress, cellular response to fluid shear stress, potassium ion transmembrane transport,  determination of liver left/right asymmetry, metanephric mesenchyme development, mesonephric tubule development, mesonephric duct development, metanephric smooth muscle tissue development, metanephric cortex development, metanephric ascending thin limb development, metanephric cortical collecting duct development, metanephric distal tubule development, metanephric S-shaped body morphogenesis, regulation of calcium ion import, inorganic cation transmembrane transport, cell-cell signaling by wnt, negative regulation of G1/S transition of mitotic cell cycle, kidney development, embryonic placenta development, embryonic placenta development, detection of nodal flow, monoatomic ion transport, monoatomic cation transport, potassium ion transport, intracellular calcium ion homeostasis, cell-matrix adhesion, positive regulation of cytosolic calcium ion concentration, receptor signaling pathway via JAK-STAT, determination of left/right symmetry, negative regulation of cell population proliferation, positive regulation of gene expression, calcium-mediated signaling, monoatomic ion transmembrane transport, cellular response to reactive oxygen species, regulation of monoatomic ion transmembrane transport, response to stimulus, detection of mechanical stimulus, centrosome duplication, establishment of localization in cell, negative regulation of ryanodine-sensitive calcium-release channel activity, negative regulation of ryanodine-sensitive calcium-release channel activity, placenta blood vessel development, placenta blood vessel development, renal tubule morphogenesis, renal tubule morphogenesis, cellular response to calcium ion, cellular response to calcium ion | 2 | miR-20a-5p, miR-106b-5p |
| *SOGA1* | Chromosome segregation, cell cycle, insulin receptor signaling pathway, regulation of autophagy, negative regulation of gluconeogenesis, cell division | 2 | miR-30b-3p, miR-149-3p |
| *SYNGAP1* | Signal transduction. Ras protein signal transduction, pattern specification process, axonogenesis, visual learning, dendrite development, regulation of GTPase activity, receptor clustering   regulation of MAPK cascade, negative regulation of neuron apoptotic process, positive regulation of GTPase activity, negative regulation of Ras protein signal transduction, regulation of synaptic plasticity, regulation of long-term neuronal synaptic plasticity, negative regulation of axonogenesis, regulation of synapse structure or activity, modulation of chemical synaptic transmission, neuron apoptotic process, maintenance of postsynaptic specialization structure | 2 | miR-92a-2-5p, miR-149-3p |
| *ZFYVE26* | Double-strand break repair via homologous recombination, cytokinesis, lysosome organization, regulation of cytokinesis, autophagosome organization, mitotic cytokinesis, DNA repair, DNA damage response, cell cycle, cell division | 2 | miR-20a-5p, miR-106b-5p |
